# Cost-effectiveness of public health strategies for COVID-19 epidemic control in South Africa: a microsimulation modelling study

**DOI:** 10.1101/2020.06.29.20140111

**Authors:** Krishna P. Reddy, Fatma M. Shebl, Julia H. A. Foote, Guy Harling, Justine A. Scott, Christopher Panella, Kieran P. Fitzmaurice, Clare Flanagan, Emily P. Hyle, Anne M. Neilan, Amir M. Mohareb, Linda-Gail Bekker, Richard J. Lessells, Andrea L. Ciaranello, Robin Wood, Elena Losina, Kenneth A. Freedberg, Pooyan Kazemian, Mark J. Siedner

## Abstract

**Background:** Healthcare resource constraints in low and middle-income countries necessitate selection of cost-effective public health interventions to address COVID-19.

**Methods:** We developed a dynamic COVID-19 microsimulation model to evaluate clinical and economic outcomes and cost-effectiveness of epidemic control strategies in KwaZulu-Natal, South Africa. Interventions assessed were Healthcare Testing (HT), where diagnostic testing is performed only for those presenting to healthcare centres; Contact Tracing (CT) in households of cases; Isolation Centres (IC), for cases not requiring hospitalisation; community health worker-led Mass Symptom Screening and molecular testing for symptomatic individuals (MS); and Quarantine Centres (QC), for household contacts who test negative. Given uncertainties about epidemic dynamics in South Africa, we evaluated two main epidemic scenarios over 360 days, with effective reproduction numbers (R_e_) of 1·5 and 1·2. We compared *HT, HT+CT, HT+CT+IC, HT+CT+IC+MS, HT+CT+IC+QC*, and *HT+CT+IC+MS+QC*, considering strategies with incremental cost-effectiveness ratio (ICER) <US$3,250/year-of-life saved (YLS) cost-effective. In sensitivity analyses, we varied R_e_, molecular testing sensitivity, and efficacies and costs of interventions.

**Findings:** With R_e_ 1·5, *HT* resulted in the most COVID-19 deaths over 360 days. Compared with *HT, HT+CT+IC+MS+QC* reduced mortality by 94%, increased costs by 33%, and was cost-effective (ICER $340/YLS). In settings where quarantine centres cannot be implemented, *HT+CT+IC+MS* was cost-effective compared with *HT* (ICER $590/YLS). With R_e_ 1·2, *HT+CT+IC+QC* was the least costly strategy, and no other strategy was cost-effective. *HT+CT+IC+MS+QC* was cost-effective in many sensitivity analyses; notable exceptions were when R_e_ was 2·6 and when efficacies of ICs and QCs for transmission reduction were reduced.

**Interpretation:** In South Africa, strategies involving household contact tracing, isolation, mass symptom screening, and quarantining household contacts who test negative would substantially reduce COVID-19 mortality and be cost-effective. The optimal combination of interventions depends on epidemic growth characteristics and practical implementation considerations.

**Funding:** National Institutes of Health, Royal Society, Wellcome Trust

## INTRODUCTION

By early September 2020, 16 countries in sub-Saharan Africa (SSA) had reported over 10,000 COVID-19 cases.^1^ High urban density, limited opportunities for physical distancing, and poor access to hygiene interventions raise the risk of severe epidemics in the region.^2^ The existing public health infrastructure for epidemic response in SSA is also of concern: testing capacity, surveillance infrastructure, isolation facilities, and intensive care (ICU) services are sparse.^3,4^

Low and middle-income countries (LMICs) are implementing epidemic control programs. The World Health Organization (WHO) promotes establishment of disease surveillance platforms, contact tracing, and isolation facilities.^5^ Epidemiologic models of these interventions have generally suggested that their efficacy depends on intervention adherence and transmission dynamics.^6,7^ Yet few studies have included resource costs to examine their cost-effectiveness and feasibility. Limitations in human resources, public health financing, and healthcare facility availability necessitate particular attention to these issues in LMICs.

We used a dynamic microsimulation model to compare medical outcomes and costs for a range of COVID-19 control measures in KwaZulu-Natal, South Africa. Our objective was to inform policy decision making by projecting clinical and economic outcomes, cost-effectiveness, and budget impact of alternative control strategies, focusing on those proposed or currently in use in South Africa. Though the first wave of diagnosed COVID-19 cases in South Africa peaked in July 2020, this analysis can inform preparation for or response to a resurgence or subsequent waves.

## METHODS

### Analytic Overview

We developed the Clinical and Economic Analysis of COVID Interventions (CEACOV) dynamic state-transition Monte Carlo microsimulation model to reflect COVID-19 natural history, diagnosis, and treatment. We compared six public health intervention strategies (figure S1). In all strategies: (a) testing consists of polymerase chain reaction (PCR) for severe acute respiratory syndrome coronavirus 2 (SARS-CoV-2) on a nasopharyngeal specimen; (b) those awaiting test results are instructed to self-isolate; (c) those severely ill (with dyspnoea and/or hypoxemia), regardless of test result, are admitted to hospital until hospital capacity is reached; those with a negative test result are advised to practice physical distancing and hand hygiene; (e) those with an initial negative test result can present for repeat testing if they develop new or worsening symptoms; (f) those not initially admitted to hospital can be admitted later if they develop severe illness. Unique characteristics of each modelled strategy are:

1. *Healthcare Testing (HT)*: Approximately 30% of people with mild/moderate COVID-19-like symptoms and all with severe symptoms self-present to a healthcare centre for testing. Those with a positive result and not severely ill are instructed to self-isolate at home.
2. *Contact Tracing (HT+CT)*: In addition to *HT*, household contacts of COVID-19 cases are tested. Those with a positive result are instructed to self-isolate at home.
3. *Contact Tracing + Isolation Centre (HT+CT+IC)*: In addition to *HT+CT*, cases who are not severely ill are referred to an isolation centre (IC) offering food, shelter, and basic medical care without supplemental oxygen. They are discharged after 14 days.
4. *Contact Tracing + Isolation Centre + Mass Symptom Screening (HT+CT+IC+MS)*: In addition to *HT+CT+IC*, community healthcare workers screen the entire population for COVID-19 symptoms every 6 months and refer those with symptoms for PCR testing. Individuals with a positive PCR test but not severely ill are referred to an IC. As a frame of reference, epidemic control measures in South Africa in June 2020 included combinations of HT, CT, IC, and MS.
5. *Contact Tracing + Isolation Centre + Quarantine Centre (HT+CT+IC+QC)*: In addition to *HT+CT+IC*, household contacts with a negative test result who cannot safely quarantine at home are referred to a quarantine centre (QC) where they receive food and shelter. They are discharged after 14 days, unless they develop COVID-19-like symptoms, in which case they are referred to ICs or hospitals, as available.
6. *Contact Tracing + Isolation Centre + Mass Symptom Screening + Quarantine Centre (HT+CT+IC+MS+QC)*: This is a combination of all measures described above.

Starting with SARS-CoV-2 infection prevalence of 0·1%, we simulated COVID-19-specific outcomes over 360 days, including daily and cumulative infections (detected and undetected), deaths, resource utilization, and healthcare costs from the health sector perspective without discounting. Outside the model, we calculated the average lifetime years-of-life saved (YLS) from each averted COVID-19 death during the 360-day model horizon, which equated to 16·8 life-years (appendix p.5-6). The primary outcome was the incremental cost-effectiveness ratio (ICER), the difference in healthcare costs (2019 US dollars [US$]) divided by the difference in life-years between strategies. We did not include costs beyond the 360-day model horizon. Average non-HIV public health expenditures in South Africa are approximately $600/year per capita;^8,9^ including those annual costs over a lifetime yields a lifetime ICER approaching $600/YLS. Therefore, our ICER estimates include healthcare costs during the 360-day model horizon and YLS over a lifetime from averted COVID-19 deaths during the 360-day model horizon. Recognizing no established threshold, we judged an ICER less than $3,250/YLS cost-effective, based on an opportunity cost approach (appendix p.2).^10^

### Model Structure

#### Health States and Natural History

At simulation initiation, each individual is either susceptible to, or infected with, SARS-CoV-2 according to age-stratified probabilities (0-19, 20-59, ≥60 years). Once infected, an individual transitions to the pre-infectious latency state. Each individual faces an age-dependent probability of developing asymptomatic, mild/moderate, severe, or critical disease (appendix p.2, table S1, figure S2). Those with critical disease face daily probabilities of death. If they survive, they pass through a recuperation state (remaining infectious) before going to the recovery state. Those in other disease states can transition directly to the recovery state. “Recovered” individuals pose no risk of transmission and are assumed immune from repeat infection for the simulation duration. All simulated individuals advance through the model simultaneously to capture infection transmission dynamics. To validate our natural history assumptions, we compared model-projected COVID-19 deaths with those reported in KwaZulu-Natal (appendix p.4).

#### Transmission

Individuals in asymptomatic, mild/moderate, severe, critical, or recuperation states of COVID-19 may transmit infection to susceptible individuals at state-dependent daily rates. The number of daily infections is a function of the proportion of susceptible people in the population, the distribution of disease states among those with COVID-19, and interventions that influence transmission (appendix p.3).

#### Testing and Interventions

PCR testing specifications include sensitivity, specificity, time from testing to result, and cost. Interventions influence testing probability (e.g., CT and MS), infection transmission rate (e.g., IC and QC), and costs.

#### Resource Utilization

Individuals with severe or critical disease are referred to hospitals and ICUs, respectively. If those resources are not available, the individual receives the next lower available intervention, which is associated with a different mortality risk and cost (e.g., if a person needs ICU care when no ICU beds are available, s/he receives non-ICU hospital care).

### Model Calibration

We populated CEACOV with COVID-19 natural history data from published literature (table 1). We used estimates of the basic reproduction number (R_0_) and viral shedding duration in various disease states to calculate transmission rates. We then calibrated transmission rates to construct an effective reproduction number (R_e_) corresponding to South African estimates in May 2020, after implementation of physical distancing and lockdown policies (appendix p.4).^11^

**Table 1.** Input parameters for a model-based analysis of COVID-19 intervention strategies in KwaZulu-Natal, South Africa.

| Parameter | Base Case Value (Range) | Source |
| --- | --- | --- |
| **Cohort Characteristics** |  |  |
| Age distribution, % |  | 12 |
| 0-19y | 40.26 |  |
| 20-59y | 51.48 |  |
| ≥60y | 8.26 |  |
| **Natural History** |  |  |
| Proportion in each health state at model start, % |  | Asm. |
| Susceptible | 99.900 |  |
| Infected |  |  |
| Pre-infectious latency | 0.030 |  |
| Asymptomatic | 0.030 |  |
| Mild/moderate disease | 0.030 |  |
| Severe disease | 0.005 |  |
| Critical disease | 0.005 |  |
| Recuperation after critical disease | 0.000 |  |
| Recovered | 0.000 |  |
| **Transmission** |  |  |
| Probability of onward transmission, daily, stratified by health state* |  | See Appendix |
| Asymptomatic | 0.1556 |  |
| Mild/moderate disease | 0.1266 |  |
| Severe disease | 0.0088 |  |
| Critical disease | 0.0070 |  |
| Recuperation after critical disease | 0.0088 |  |
| Effective reproductive number ($R_e$, range) | 1.5 (1.1-2.6) | 11 |

| **PCR Testing** |  |  |
| --- | --- | --- |
| Sensitivity<sup>†</sup>, nasopharyngeal specimen, % (range) | 70 (50-90) | 30,31 |
| Specificity, nasopharyngeal specimen, % | 100 | Asm. |
| Cost, 2019 US\$ (range) | 26 (13-52) | AHRI |
| Time to result return and action, days (range) | 5 (1-7) | AHRI |
| **Resource Utilization** |  |  |
| Resources available per 11,000,000 people, n |  |  |
| Hospital beds | 26,220 | 14 |
| ICU beds | 748 | 14 |
| Isolation centre beds | As needed, no capacity limitation | Asm. |
| Quarantine centre beds | As needed, no capacity limitation | Asm. |
| Cost, per person, 2019 US\$ (range) | | |
| Hospital bed, daily | 165 (83-330) | 32 |
| ICU bed, daily | 2,059 (1,030-4,118) | 33 |
| Contact tracing/mass symptom screen, per instance | 3 (2-6) | AHRI |
| Isolation centre bed, daily | 44 (22-88) | AHRI |
| Quarantine centre bed, daily | 37 (19-74) | AHRI |

COVID-19: coronavirus disease 2019. y: years. Asm.: assumption. PCR: polymerase chain reaction. US\$: United States dollars. ICU: intensive care unit. AHRI: Africa Health Research Institute (KwaZulu-Natal, South Africa; personal communication).
Values indicated are those applied in the base case analyses or, in parentheses, the ranges evaluated in sensitivity analysis.
\*These values reflect transmission probabilities in a scenario in which $R_e$ is 1.5.
<sup>†</sup>Test sensitivity does not vary by disease stage, except that it is 0% in the pre-infectious latency phase.

### Input Parameters

#### Cohort Characteristics

We defined cohort demographic characteristics using 2019 population estimates (table 1).^12^ In KwaZulu-Natal, 40.26% were aged 0-19 years, 51.48% were 20-59 years, and 8.26% were over 60 years. Day 0 of the model represents a provincial 0·1% prevalence (approximately 11,000 individuals) of active SARS-CoV-2 infection, with the remainder of the population susceptible to infection.

#### Natural History

For those newly infected with SARS-CoV-2, average pre-infectious latency was 2·6 days. Table S1 indicates duration in each state, age-dependent probability of developing severe or critical disease, and age-dependent mortality for those with critical disease.

#### Transmission

We stratified transmission rates by disease state (table 1). We adjusted transmission rates to reflect R_e_=1·5.^11^ Given uncertainty over R_e_, both in the past and future, we also simulated alternative epidemic growth scenarios with lower (1·1 or 1·2) or higher (2·6) R_e_ (appendix p.4).

#### Testing and Interventions

In the base case, we assumed a 70% sensitivity, 100% specificity, and five days to PCR result return and action across all active infection states.^13^ We defined the probability of undergoing testing based on the health state and intervention strategy in place (table S2, appendix p.7). Given limited data about the precise efficacy of each intervention for reducing SARS-CoV-2 transmission rates (e.g., IC), we made assumptions about efficacies and varied them in sensitivity analysis. Ongoing transmission after diagnosis was reduced by 50% from HT alone and by 95% when HT was combined with ICs or QCs (table S2).

#### Resource Utilization and Costs

The maximum capacity of hospital and ICU beds was 26,220 and 748 per 11 million people, as reported for KwaZulu-Natal (table 1).^14^ We assumed that IC and QC beds were available to all who needed them. We applied costs of PCR testing, contact tracing, and mass symptom screening, as well as daily costs of hospitalisation, ICU stay, and IC/QC stays based on published estimates and/or cost quotes obtained in KwaZulu-Natal (appendix p.6).

### Resource Utilization and Budget Impact Analysis

We conducted resource utilization and budget impact analysis from a combined public/private health sector perspective for KwaZulu-Natal (population 11 million). We projected the total resources, including testing, hospital/ICU beds, and IC/QC beds, that would be used in each intervention strategy. IC/QC beds are offered to those who meet criteria, and we assumed in the base case that all offered would be used. In budget impact analysis, we projected total and component healthcare costs associated with each strategy over 360 days and compared them with the 2019 KwaZulu-Natal Department of Health budget of $3·12 billion.^15^ Because ICU care is relatively expensive and mostly in the private sector, we also considered costs exclusive of ICU care.

### Sensitivity Analysis

We conducted one-way sensitivity analysis by varying key parameters across plausible ranges to determine the impact on clinical and cost projections (table 1, table S2). To extrapolate to other settings, we limited hospital and ICU bed availability to the average numbers in SSA countries (22,275 and 371 per 11 million people).^16^ We performed multi-way sensitivity analysis in which we varied parameters influential in one-way sensitivity analysis, including reducing IC/QC efficacy and costs to reflect the impact of home-based isolation and quarantine strategies.

## RESULTS

For an epidemic with R_e_=1·5, we projected that *HT* would result in the most COVID-19 infections and deaths, most life-years lost, and lowest costs over 360 days (table 2, figures S3-S4). *HT+CT+IC+MS+QC* provided the greatest clinical benefit and was cost-effective (ICER $340/YLS) (figure 1). *HT+CT+IC+MS+QC* decreased life-years lost by 94% and increased costs by 33% compared with *HT*. All other strategies resulted in higher costs while providing less clinical benefit than *HT+CT+IC+MS+QC*. In settings where quarantine centres cannot be implemented, *HT+CT+IC+MS* was the cost-effective strategy (ICER $590/YLS compared with *HT*).

**Table 2.**
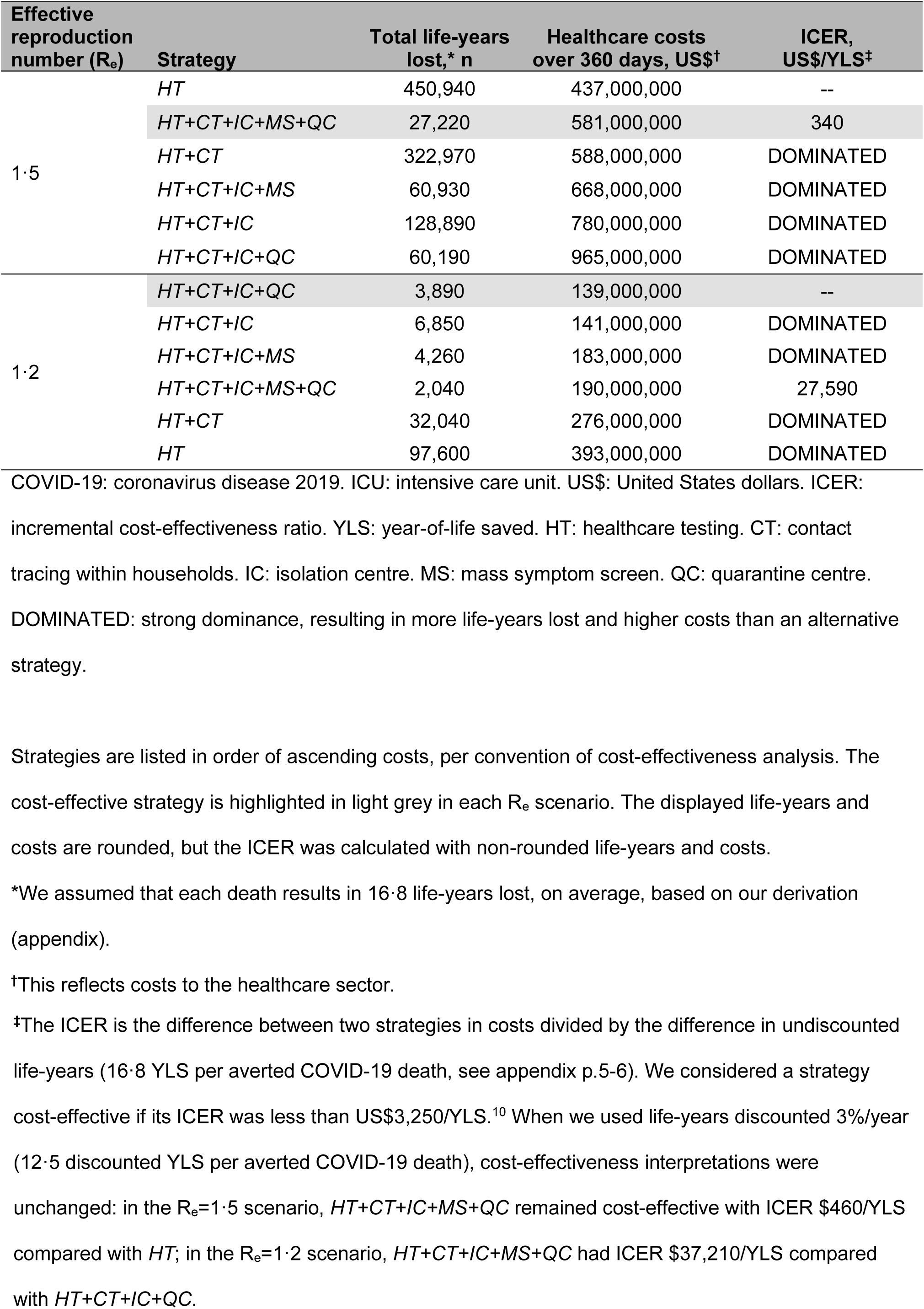
Model-projected life-years lost, healthcare costs, and cost-effectiveness of COVID-19 intervention strategies in KwaZulu-Natal, South Africa.

| Effective reproduction number ($R_e$) | Strategy | Total life-years lost,* n | Healthcare costs over 360 days, US\$<sup>†</sup> | ICER, US\$/YLS<sup>‡</sup> |
| --- | --- | --- | --- | --- |
| 1.5 | HT | 450,940 | 437,000,000 | -- |
|  | HT+CT+IC+MS+QC | 27,220 | 581,000,000 | 340 |
|  | HT+CT | 322,970 | 588,000,000 | DOMINATED |
|  | HT+CT+IC+MS | 60,930 | 668,000,000 | DOMINATED |
|  | HT+CT+IC | 128,890 | 780,000,000 | DOMINATED |
|  | HT+CT+IC+QC | 60,190 | 965,000,000 | DOMINATED |
| 1.2 | HT+CT+IC+QC | 3,890 | 139,000,000 | -- |
|  | HT+CT+IC | 6,850 | 141,000,000 | DOMINATED |
|  | HT+CT+IC+MS | 4,260 | 183,000,000 | DOMINATED |
|  | HT+CT+IC+MS+QC | 2,040 | 190,000,000 | 27,590 |
|  | HT+CT | 32,040 | 276,000,000 | DOMINATED |
|  | HT | 97,600 | 393,000,000 | DOMINATED |

COVID-19: coronavirus disease 2019. ICU: intensive care unit. US\$: United States dollars. ICER: incremental cost-effectiveness ratio. YLS: year-of-life saved. HT: healthcare testing. CT: contact tracing within households. IC: isolation centre. MS: mass symptom screen. QC: quarantine centre. DOMINATED: strong dominance, resulting in more life-years lost and higher costs than an alternative strategy.
Strategies are listed in order of ascending costs, per convention of cost-effectiveness analysis. The cost-effective strategy is highlighted in light grey in each $R_e$ scenario. The displayed life-years and costs are rounded, but the ICER was calculated with non-rounded life-years and costs.
\*We assumed that each death results in 16.8 life-years lost, on average, based on our derivation (appendix).
<sup>†</sup>This reflects costs to the healthcare sector.
\*The ICER is the difference between two strategies in costs divided by the difference in undiscounted life-years (16.8 YLS per averted COVID-19 death, see appendix p.5-6). We considered a strategy cost-effective if its ICER was less than US\$3,250/YLS.<sup>10</sup> When we used life-years discounted 3%/year (12.5 discounted YLS per averted COVID-19 death), cost-effectiveness interpretations were unchanged: in the $R_e=1.5$ scenario, $HT+CT+IC+MS+QC$ remained cost-effective with ICER \$460/YLS compared with $HT$ ; in the $R_e=1.2$ scenario, $HT+CT+IC+MS+QC$ had ICER \$37,210/YLS compared with $HT+CT+IC+QC$ .

**Figure 1.**
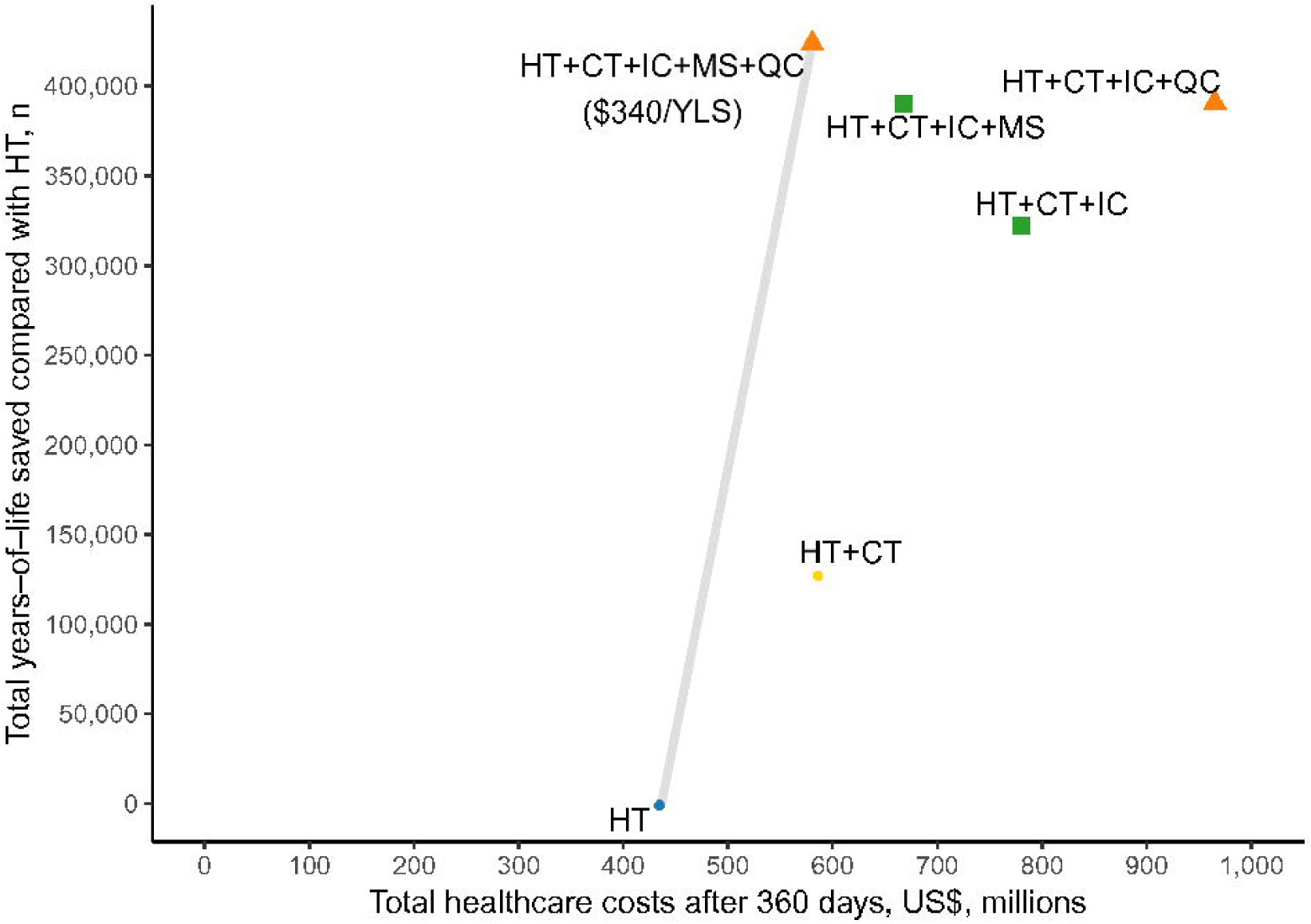
Cost-effectiveness efficiency frontier: COVID-19 intervention strategies in KwaZulu-Natal, South Africa. COVID-19: coronavirus disease 2019. HT: healthcare testing. CT: contact tracing within households. IC: isolation centre. MS: mass symptom screen. QC: quarantine centre. Model results are shown for an effective reproduction number of 1·5. Strategies that are below the line are dominated – i.e., an inefficient use of resources compared with other strategies. For non-dominated strategies, ICERs are shown below the strategy label.

With R_e_=1·2, *HT+CT+IC+QC* was cost-saving compared with *HT* (table 2). *HT+CT+IC+MS+QC* resulted in 48% fewer life-years lost but was not cost-effective (ICER $27,590/YLS) compared with *HT+CT+IC+QC. HT+CT+IC* was the least costly strategy in settings where quarantine centres cannot be implemented, and other strategies were not cost-effective compared with *HT+CT+IC*.

Regarding resource utilization, with R_e_=1·5, *HT* resulted in the highest peak daily use of hospital beds (table 3). Compared with *HT, HT+CT+IC+MS+QC* increased cumulative PCR test usage by 2.6 times (though with lower peak daily PCR use) and reduced peak daily hospital bed use by 86% (due to fewer cumulative infections), while requiring 12,380 IC beds and 18,140 QC beds at peak daily use. Only the *HT+CT+IC+MS, HT+CT+IC+QC*, and *HT+CT+IC+MS+QC* strategies maintained peak daily ICU bed demand below provincial capacity.

**Table 3.** Model-projected resource utilization of COVID-19 intervention strategies in KwaZulu-Natal, South Africa.

| Effective reproduction number ($R_e$) | Strategy | Cumulative PCR tests performed over 360 days, n | Peak daily resource use, n | | | | |
| --- | --- | --- | --- | --- | --- | --- | --- |
|  |  |  | PCR tests | Hospital beds (non-ICU) | ICU beds* | Isolation centre beds | Quarantine centre beds |
| 1.5 | *HT* | 1,527,450 | 14,820 | 4,690 | 748 | -- | -- |
|  | *HT+CT+IC+MS+QC* | 3,904,230 | 12,900 | 640 | 341 | 12,380 | 18,140 |
|  | *HT+CT* | 5,951,180 | 31,050 | 3,440 | 748 | -- | -- |
|  | *HT+CT+IC+MS* | 4,639,280 | 16,930 | 1,320 | 715 | 21,260 | -- |
|  | *HT+CT+IC* | 4,904,010 | 19,340 | 1,930 | 748 | 30,510 | -- |
|  | *HT+CT+IC+QC* | 4,478,770 | 16,710 | 1,380 | 737 | 26,710 | 39,470 |
| 1.2 | *HT+CT+IC+QC* | 2,963,280 | 9,870 | 590 | 363 | 1,840 | 3,110 |
|  | *HT+CT+IC* | 3,025,260 | 9,870 | 590 | 363 | 1,620 | -- |
|  | *HT+CT+IC+MS* | 3,159,950 | 10,520 | 570 | 396 | 1,510 | -- |
|  | *HT+CT+IC+MS+QC* | 3,120,800 | 10,520 | 570 | 396 | 1,860 | 3,480 |
|  | *HT+CT* | 3,647,570 | 12,450 | 770 | 506 | -- | -- |
|  | *HT* | 766,140 | 4,440 | 1,680 | 748 | -- | -- |

COVID-19: coronavirus disease 2019. PCR: polymerase chain reaction. ICU: intensive care unit. HT: healthcare testing. CT: contact tracing within households. IC: isolation centre. MS: mass symptom screen. QC: quarantine centre.
Strategies are listed in order of ascending costs as indicated in table 2, per convention of cost-effectiveness analysis. The cost-effective strategy is highlighted in light grey in each $R_e$ scenario.
\*The total number of available ICU beds was 748.

With R_e_=1·2, compared with *HT, HT+CT+IC+MS+QC* increased cumulative PCR test usage by 4.1 times and reduced peak daily hospital bed use by 66%, while requiring 1,860 IC beds and 3,480 QC beds at peak daily use. All strategies except *HT* maintained peak daily ICU bed demand below capacity.

Over 360 days, for an epidemic with R_e_=1·5, PCR testing contributed 9-27% to overall costs, depending on the strategy (figure 2). In strategies with QCs, these centres contributed 26-30% to overall costs. In strategies without QCs, ICU care was the largest contributor to costs, ranging from 38-71%. Costs exclusive of ICU care were $125 million (*HT*), $413 million (*HT+CT+IC+MS*), and $461 million (*HT+CT+IC+MS+QC*), reflecting approximately 4%, 13%, and 15% of the 2019 KwaZulu-Natal Department of Health budget. CT and MS together contributed ≤10% and ICs contributed 22-31% to overall costs in strategies in which they were used. For an epidemic with R_e_=1·2, costs exclusive of ICU care were $71 million (*HT*), $159 million (*HT+CT+IC+MS*), and $167 million (*HT+CT+IC+MS+QC*), reflecting 2%, 5%, and 5% of the 2019 KwaZulu-Natal Department of Health budget.

**Figure 2.**
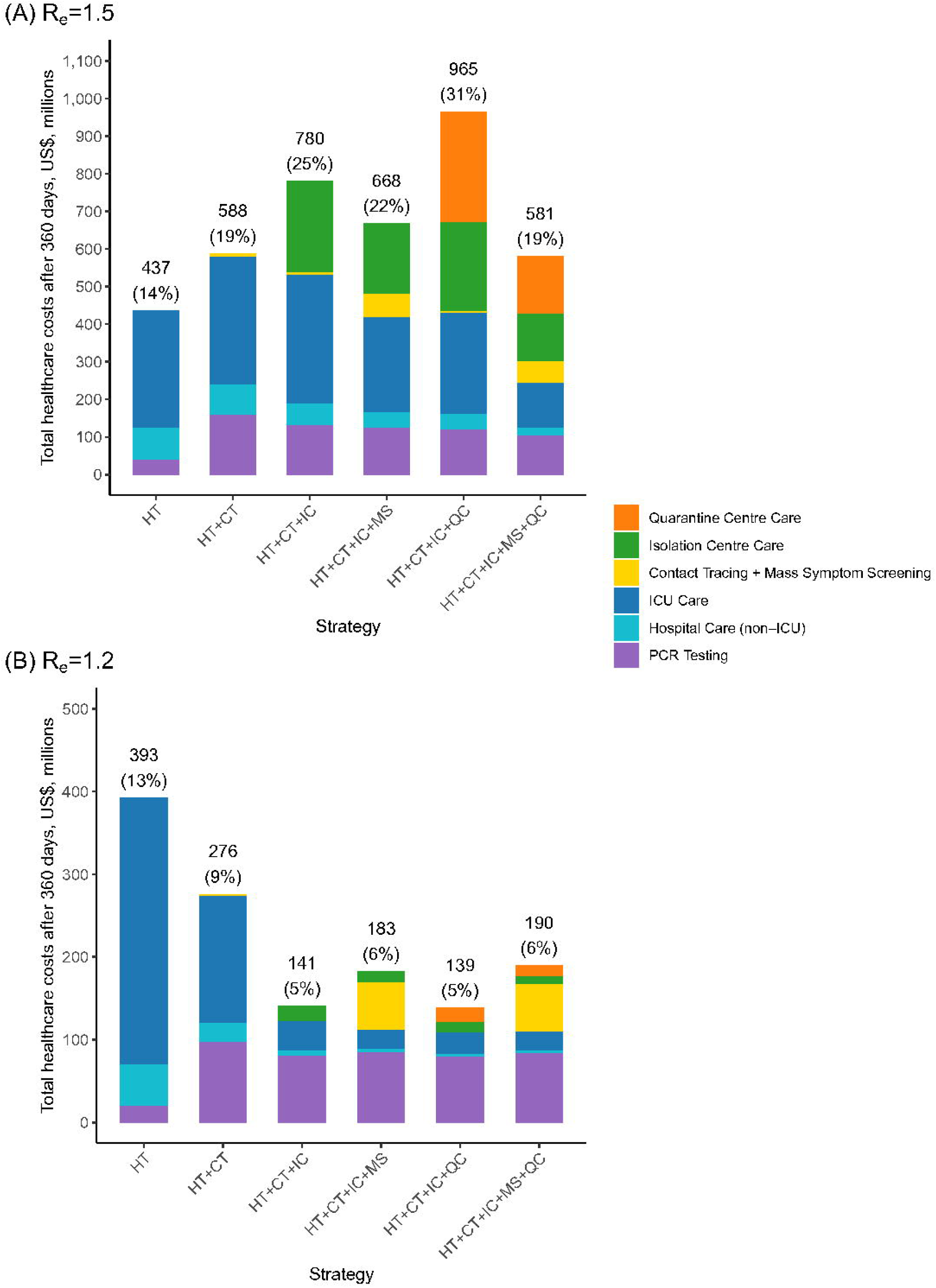
Budget impact analysis: contributors to healthcare costs of COVID-19 intervention strategies in KwaZulu-Natal, South Africa. SARS-CoV-2: severe acute respiratory syndrome coronavirus 2. COVID-19: coronavirus disease 2019. R_e_: effective reproduction number. HT: healthcare testing. CT: contact tracing within households. IC: isolation centre. MS: mass symptom screen. QC: quarantine centre. Panel A shows results for an epidemic with R_e_=1·5, and Panel B shows results for an epidemic with R_e_=1·2. The figures show the total and component COVID-19-related healthcare costs, from a health sector perspective, associated with different intervention strategies when applied to the entire KwaZulu-Natal population of 11 million people. The costs are derived from model-generated results. Percentages in parentheses represent the proportion of the 2019 KwaZulu-Natal Department of Health budget.

In sensitivity analyses, results were similar to the base case (i.e., *HT+CT+IC+MS+QC* remained cost-effective) when varying costs of CT and MS (table S3) and hospitalisation (table S4); varying PCR sensitivity and time to result (table S5) and PCR cost (table S6); and varying availability of hospital and ICU beds (table S7). When PCR sensitivity increased to 90%, both *HT+CT+IC+MS* (ICER $440/YLS) and *HT+CT+IC+MS+QC* (ICER $1,660/YLS) used resources efficiently.

Conversely, our projected ICERs changed meaningfully in a model with R_e_=2·6 – resource requirements increased substantially, making *HT+CT+IC+MS+QC* no longer cost-effective (ICER $25,040/YLS), and all strategies had ICERs above our cost-effectiveness threshold compared with *HT* (table S8). The pattern of results with R_e_=1·1 was similar to that with R_e_=1·2 (table S8). When the efficacies of CT and MS to detect infections were halved from the base case values, *HT+CT+IC+MS+QC* was no longer cost-effective (ICER $5,930/YLS) (table S9). When the efficacy of ICs/QCs for transmission reduction was decreased from 95% to 75%, *HT+CT+IC+MS+QC* was not cost-effective ($12,490/YLS) (table S10). When the IC/QC costs decreased, *HT+CT+IC+MS+QC* became more favourable in terms of cost-effectiveness, and it remained cost-effective when IC/QC costs were double those of the base case (table S11).

In a multi-way sensitivity analysis that varied CT/MS efficacy and reduced IC/QC efficacy and cost to assess lower-cost but potentially lower-efficacy home-based IC and QC programs, *HT+CT+IC+MS+QC* or *HT+CT+IC+QC* were cost-effective in nearly all scenarios in which CT/MS efficacy for case detection was double that of the base case efficacy (figure S5). When CT/MS efficacy for case detection was half that of the base case efficacy, strategies involving quarantine centres were less likely to be cost-effective. If quarantine centers were not an option, then *HT+CT+IC+MS* was cost-effective in most scenarios (figure S6).

## DISCUSSION

Public health strategies combining contact tracing, isolation of those with confirmed COVID-19, community-based mass symptom screening, and quarantine of household contacts of confirmed cases will substantially reduce infections, hospitalisations, and deaths while efficiently using healthcare resources in KwaZulu-Natal, South Africa. We estimate that a strategy combining all interventions would cost an additional $340 per year-of-life saved, which compares favourably with the cost-effectiveness of many established public health interventions in South Africa, including tuberculosis diagnostic testing^17^ and cervical cancer screening.^18^ In scenarios in which implementation of quarantine centres cannot be accomplished, a strategy of contact tracing, isolation centres, and mass symptom screening would be cost-effective.

Notably, the cost-effectiveness of strategies was sensitive to epidemic growth conditions. We conducted sensitivity analyses intended to generalize to other settings with resource constraints, to epidemics at varying degrees of acceleration (including published estimates in South Africa^11,19^), and with varying intervention costs.^20^ With low epidemic growth (R_e_ 1·1-1·2), *HT+CT+IC+QC* was the optimal strategy; QCs remained cost-effective but adding MS was not cost-effective. With high epidemic growth (R_e_ 2·6), when the epidemic outpaced control measures and costs increased substantially, no combination of the modelled interventions was cost-effective compared with *HT* alone.

Our model parameters and specifications were selected for their relevance to LMICs. Our estimates are based on the population structure of KwaZulu-Natal, with a median age of 25 years (compared with 38 years in the USA), and thus are likely to reflect epidemic scenarios in LMICs with similarly young age structures. We chose intervention scenarios based on prior work supporting their efficacy for epidemic control, WHO recommendations, and particular relevance to settings with limitations in formal healthcare infrastructure.^5–7^ We did not limit the PCR testing availability – so that the total number of tests needed and associated costs could be estimated – and peak PCR use reached approximately 10,000-15,000 tests/day in the optimal strategies, marginally above established capacity in KwaZulu-Natal during the recent surge.^21^ We specified the model to reflect the number of available hospital and ICU beds in KwaZulu-Natal,^14^ and results were similar when we further restricted bed availability to that elsewhere in SSA.^16^ Contact tracing and community-based screening have been frequently used for case-finding in LMICs.^22^ Many SSA countries are thus theoretically poised to implement such interventions through established networks of community health workers. Finally, isolation centres, which are likely to require the greatest investment in new infrastructure, have been implemented successfully in response to Ebola epidemics in West Africa and the Democratic Republic of Congo, where healthcare resources are among the lowest in the world.^23^ South Africa has rapidly implemented and expanded COVID-19 related services in recent months, but further scale-up would be required to meet demand in some of our modelled scenarios.^21,24^Isolation centres in our model are designed as housing facilities for people with confirmed COVID-19 who do not require hospital-level care but cannot safely isolate at home. We estimated that their use reduces ongoing transmission after a confirmed diagnosis from 50% (in the *HT* strategy) to 5%. They are likely to be most effective in areas with high household density and limited capacity for in-home isolation, as is the case for many urban centres in SSA. Quarantine centres, which include optional housing for contacts who test negative and cannot safely distance during the latency period, have also been proposed for interrupting epidemic spread and were implemented in the early phases of the COVID-19 response in China. They were effective in our model at reducing the deleterious impact of the epidemic and were cost-effective in many modelled scenarios.

Importantly, there are critical social and human rights considerations to implementation of isolation and quarantine in many settings, due to trade-offs between public health benefits and civil liberties.^25^ In our model, both interventions are provided optionally for those who cannot do so safely at home, but we conservatively included costs to reflect needs should they be used. We also considered the use of home-based isolation and quarantine in a multi-way analysis that reduced efficacies and costs of both. We found that isolation and quarantine remained cost-effective in some lower efficacy scenarios, particularly if their costs were also reduced. On balance, from a public health perspective, our findings support use of quarantine centres in areas with individual and community support for their use.

Our model should be interpreted within the context of several limitations. We did not account for heterogenous mixing within the population. Instead, we assumed that contact patterns were random, as commonly done in infectious disease models. We assumed that the age-adjusted prevalence of non-communicable co-morbidities in South Africa would be similar to that in in the US and that age would be the primary driver of COVID-19 outcomes as demonstrated in multiple settings.^26–28^ In line with most published studies, we conservatively assumed no modifying effect of HIV on the severity of COVID-19, though additional data are needed from HIV-endemic countries to support this.^28,29^ If the high prevalence of non-communicable diseases and/or HIV in South Africa does worsen COVID-19 outcomes compared with resource-rich settings, then the benefit of public health interventions in terms of years-of-life saved and cost-effectiveness will likely be greater than our estimates. Nonetheless, in extending projections beyond the 360-day model horizon, we accounted for South Africa-specific mortality rates in our calculations of life expectancy and years-of-life lost. It will be crucial to consider how resources and interventions implemented in response to COVID-19 will impact available resources for other regional healthcare priorities. We did not include lifetime costs of healthcare beyond COVID-19 or of sequelae among the recovered, and we did not account for impacts of COVID-19 interventions on other economic sectors. As with all modelling exercises, our estimates are determined by assumptions of input parameters. We selected COVID-19 clinical parameters based on the published literature, which are largely derived from high-income settings. Intervention efficacy estimates were hypothesized based on other model parameters, existing literature where available, or expert opinion if no data were available. Recognizing a lack of empiric data for some of these estimates, we focused our sensitivity analyses on varying those for which data was lacking. Finally, costing data were derived from the literature and direct cost estimates from local suppliers in KwaZulu-Natal and therefore might not reflect costs in other contexts nor full implementation and scale-up costs. Nonetheless, our primary findings and policy conclusions were largely consistent across a range of costing estimates.

We recommend that policymakers consider contact tracing, isolation of confirmed cases, mass symptom screening, and quarantine of household contacts of cases to address COVID-19 epidemic control efficiently. Where quarantine centres are not feasible – for example, due to budget constraints or lack of public support – a strategy that includes the other interventions would still provide clinical benefit in an economically efficient manner.

## Data Availability

The relevant data are contained within the manuscript and appendix, in published papers, or on publicly available websites.

## AUTHOR ROLES

All authors contributed substantively to this manuscript in the following ways: study and model design (all authors), data analysis (KPR, FMS, JHAF, GH, KPF, KAF, PK, MJS), interpretation of results (all authors), drafting the manuscript (KPR, MJS), critical revision of the manuscript (all authors) and final approval of submitted version (all authors).

## DATA SHARING STATEMENT

This modelling study involved the use of published or publicly available data. The data used and the sources are described in the manuscript and appendix. No primary data were collected for this study. Model flowcharts are in the appendix.

## ACKNOWLEDGMENTS

This work was supported by the National Institutes of Health [R37 AI058736, K24 AR057827, and T32 AI007433] and by a fellowship from the Royal Society and Wellcome Trust [210479/Z/18/Z].

The funding sources had no role in the study design, data collection, data analysis, data interpretation, writing of the manuscript, or in the decision to submit the manuscript for publication. The content is solely the responsibility of the authors and does not necessarily represent the official views of the funding sources.

The authors thank Nicole McCann for technical assistance.

## REFERENCES

1 Xu B, Kraemer MUG, Xu B, et al. Open access epidemiological data from the COVID-19 outbreak. Lancet Infect Dis 2020; 20: 534.

2 Johnstone-Robertson SP, Mark D, Morrow C, et al. Social mixing patterns within a South African township community: implications for respiratory disease transmission and control. Am J Epidemiol 2011; 174: 1246–55.

3 Siedner MJ, Gostin LO, Cranmer HH, Kraemer JD. Strengthening the detection of and early response to public health emergencies: lessons from the West African Ebola epidemic. PLoS Med 2015; 12: e1001804.

4 Gilbert M, Pullano G, Pinotti F, et al. Preparedness and vulnerability of African countries against importations of COVID-19: a modelling study. Lancet 2020; 395: 871–7.

5 World Health Organization. COVID-19 Strategic Preparedness and Response Plan: Operational planning guidelines to support country preparedness and response. 2020; published online Feb 12. https://www.who.int/docs/default-source/coronaviruse/covid-19-sprp-unct-guidelines.pdf?sfvrsn=81ff43d8_4 (accessed June 24, 2020).

6 Hellewell J, Abbott S, Gimma A, et al. Feasibility of controlling COVID-19 outbreaks by isolation of cases and contacts. Lancet Glob Health 2020; 8: e488–96.

7 Peak CM, Childs LM, Grad YH, Buckee CO. Comparing nonpharmaceutical interventions for containing emerging epidemics. Proc Natl Acad Sci 2017; 114: 4023–8.

8 Basu S, Wagner RG, Sewpaul R, Reddy P, Davies J. Implications of scaling up cardiovascular disease treatment in South Africa: a microsimulation and cost-effectiveness analysis. Lancet Glob Health 2019; 7: e270–80.

9 Global Burden of Disease Health Financing Collaborator Network. Spending on health and HIV/AIDS: domestic health spending and development assistance in 188 countries, 1995-2015. Lancet 2018; 391: 1799–829.

10 Edoka IP, Stacey NK. Estimating a cost-effectiveness threshold for health care decision-making in South Africa. Health Policy Plan 2020; 35: 546–55.

11 National Institute for Communicable Diseases. The Initial and Daily COVID-19 Effective Reproductive Number (R) in South Africa. 2020; published online May 27. https://www.nicd.ac.za/wp-content/uploads/2020/05/The-Initial-and-Daily-COVID-19-Effective-Reproductive-Number-R-in-South-Africa-002.pdf (accessed June 24, 2020).

12 Statistics South Africa. Mid-year population estimates 2019. http://www.statssa.gov.za/publications/P0302/P03022019.pdf (accessed Sept 7, 2020).

13 National Institute for Communicable Diseases. COVID-19 Testing Summary. 2020; published online May 23. https://www.nicd.ac.za/wp-content/uploads/2020/05/NICD-COVID-19-Testing-Summary_-Week-21-2020-007.pdf (accessed June 24, 2020).

14 National Department of Health, South Africa. COVID-19 Public Health Response. 2020; published online April 10. https://sacoronavirus.co.za/2020/04/11/covid-19-public-health-response/ (accessed June 24, 2020).

15 Province of KwaZulu-Natal, Department of Health. Annual Performance Plan 2019/20-2021/22. 2019. http://www.kznhealth.gov.za/app/APP-2019-20.pdf (accessed June 24, 2020).

16 Craig J, Kalanxhi E, Hauck S. National estimates of critical care capacity in 54 African countries. Public and Global Health, 2020 DOI:10.1101/2020.05.13.20100727.

17 Reddy KP, Gupta-Wright A, Fielding KL, et al. Cost-effectiveness of urine-based tuberculosis screening in hospitalised patients with HIV in Africa: a microsimulation modelling study. Lancet Glob Health 2019; 7: e200–8.

18 Goldie SJ, Gaffikin L, Goldhaber-Fiebert JD, et al. Cost-effectiveness of cervical-cancer screening in five developing countries. N Engl J Med 2005; 353: 2158–68.

19 Centre for the Mathematical Modelling of Infectious Disases, London School of Hygiene and Tropical Medicine. COVID-19 Estimates for South Africa. 2020; published online June 22. https://epiforecasts.io/covid/posts/national/south-africa/ (accessed June 24, 2020).

20 Siedner MJ, Harling G, Reynolds Z, et al. Social distancing to slow the US COVID-19 epidemic: Longitudinal pretest-posttest comparison group study. PLoS Med 2020; 17: e1003244.

21 National Institute for Communicable Diseases, South Africa. COVID-19 Testing Summary. NICD. 2020; published online March 2. https://www.nicd.ac.za/wp-content/uploads/2020/08/COVID-19-Testing-Summary-Week-34-2020.pdf (accessed Sept 2, 2020).

22 Shapiro AE, Variava E, Rakgokong MH, et al. Community-based targeted case finding for tuberculosis and HIV in household contacts of patients with tuberculosis in South Africa. Am J Respir Crit Care Med 2012; 185: 1110–6.

23 Legrand J, Grais RF, Boelle PY, Valleron AJ, Flahault A. Understanding the dynamics of Ebola epidemics. Epidemiol Infect 2007; 135: 610–21.

24 Abdool Karim SS. The South African response to the pandemic. N Engl J Med 2020; 382: e95.

25 Blendon RJ, Koonin LM, Benson JM, et al. Public response to community mitigation measures for pandemic influenza. Emerg Infect Dis 2008; 14: 778–86.

26 Lewnard JA, Liu VX, Jackson ML, et al. Incidence, clinical outcomes, and transmission dynamics of severe coronavirus disease 2019 in California and Washington: prospective cohort study. BMJ 2020; 369: m1923.

27 Docherty AB, Harrison EM, Green CA, et al. Features of 20?133 UK patients in hospital with covid-19 using the ISARIC WHO Clinical Characterisation Protocol: prospective observational cohort study. BMJ 2020; 369: m1985.

28 Boulle A, Davies M-A, Hussey H, et al. Risk factors for COVID-19 death in a population cohort study from the Western Cape Province, South Africa. Clin Infect Dis 2020; published online Aug 29. DOI:10.1093/cid/ciaa1198.

29 del Rio C. COVID-19 in Persons Living with HIV — What Do We Know Today? NEJM J Watch 2020; 2020. DOI:10.1056/nejm-jw.NA52137.

30 Yang Y, Yang M, Shen C, et al. Evaluating the accuracy of different respiratory specimens in the laboratory diagnosis and monitoring the viral shedding of 2019-nCoV infections. Infectious Diseases (except HIV/AIDS), 2020 DOI:10.1101/2020.02.11.20021493.

31 Wang W, Xu Y, Gao R, et al. Detection of SARS-CoV-2 in different types of clinical specimens. JAMA 2020; 323: 1843–4.

32 Netcare Hospitals. Netcare Tariffs. 2016. https://www.netcarehospitals.co.za/ (accessed June 10, 2020).

33 Mahomed S, Mahomed OH. Cost of intensive care services at a central hospital in South Africa. S Afr Med J 2018; 109: 35.

