## Appendix for "Cost-effectiveness of public health strategies for COVID-19 epidemic control in South Africa: a microsimulation modelling study"

**Contents**

S1. Methods: Additional Information 2
Appendix References 9
Table S1 11
Table S2 12
Table S3 13
Table S4 14
Table S5 16
Table S6 17
Table S7 18
Table S8 19
Table S9 20
Table S10 21
Table S11 22
Table S12 23
Table S13 24
Table S14 25
Table S15 26
Table S16 27
Figure S1 28
Figure S2 32
Figure S3 33
Figure S4 34
Figure S5 35
Figure S6 36

### METHODS: ADDITIONAL INFORMATION

#### S1.1. Model Structure and Analytic Overview

***Overview***

The Clinical and Economic Analysis of COVID Interventions (CEACOV) model consists of several modules that together determine individual health/disease trajectories and epidemic growth. These modules include natural history of disease, transmission, interventions including testing, and resource utilization.

Each model simulation in this analysis started with 1 million individuals. We then used the model to project outcomes over 360 days and extrapolated the results to the KwaZulu-Natal population of 11 million. The incremental cost-effectiveness ratio (ICER) was expressed in terms of undiscounted COVID-19-related healthcare costs during the 360-day model simulation period divided by undiscounted lifetime years-of-life saved (YLS) per COVID-19 death averted during the 360-day model simulation. We also considered YLS when discounted 3%/year (see Section S1.3).

There is much debate around appropriate cost-effectiveness thresholds, especially in low and middle-income countries. In this analysis, we applied an opportunity cost-based threshold for South Africa as reported by Edoka and Stacey.[1] We converted their reported threshold ($3,015) from 2015 United States dollars (USD) to 2015 South African Rand (ZAR), adjusted for inflation to obtain a value in 2019 ZAR, and subsequently converted from 2019 ZAR to 2019 USD to yield a threshold of $3,250 per year-of-life saved.[2,3]

***Health States***

CEACOV simulates individuals transitioning between the states of susceptibility to severe acute respiratory syndrome coronavirus 2 (SARS-CoV-2), infection with SARS-CoV-2 and COVID-19 disease, recovery from COVID-19, and death. Susceptible individuals face a daily probability of exposure. After being infected with SARS-CoV-2, individuals may progress through the following health states (Figure S2):

- Pre-infectious latency
- Asymptomatic (or presymptomatic) infection
- Mild/moderate disease: symptomatic
- Severe disease: dyspnoea and/or hypoxemia ideally managed in a hospital with standard supplemental oxygen but not requiring intensive care unit (ICU)
- Critical disease: ideally managed in an ICU with high-flow supplemental oxygen, non-invasive positive pressure ventilation, or invasive mechanical ventilation
- Recuperation: only for those recuperating from critical disease and improving while remaining in the hospital or other health care facility
- Recovered

Individuals in the asymptomatic infection, mild/moderate disease, and severe disease states can transition directly to the recovered state. Individuals in the critical disease state can eventually die, or transition to the recuperation state and then to the recovered state. The recovered state is an absorbing state, and recovered individuals are assumed to have full immunity to SARS-CoV-2 over the model time horizon.

***Natural History Paths***

After being infected with SARS-CoV-2, a susceptible individual first transitions to the pre-infectious latency stage. Then, the individual has an age-dependent probability of progressing along one of four “paths,” culminating in either asymptomatic infection, mild/moderate disease, severe disease, or critical disease. Before reaching a more advanced disease state, individuals must first transition through intermediate states (e.g., those destined for severe disease must first pass through the asymptomatic/presymptomatic infection state and the mild/moderate disease state) (Figure S2).

***Transmission***

In CEACOV,

Effective Transmission Rate ($R_{eff}$) = Nominal Transmission Rate ($R_{nom}$) * Transmission Multiplier

The nominal transmission rate ($R_{nom}$) is a function of the average number of susceptible persons whom an infected individual contacts per day in a fully susceptible cohort multiplied by the probability of infecting the susceptible person per contact. This nominal transmission rate captures the ratio (not the magnitude) of daily infectivity stratified by disease states in an index epidemic. In other words, it captures the ratio of 'force of transmission' across different disease states. Infected individuals do not transmit while they are in the pre-infectious latency state or in the recovered state. Individuals in other infected states can transmit SARS-CoV-2 to susceptible individuals. The effective transmission rate ($R_{eff}$) changes over time as social interventions alter the number of contacts and infectivity per contact. Thus, the magnitude of the transmission rate is adjusted using the transmission multiplier (see below). In a sense, the effective transmission rate ($R_{eff}$) in CEACOV is the effective reproductive number ($R_e$) divided by the average duration of infectivity.

Transmission multipliers are setting-specific, time-dependent adjusting factors. They roughly account for population density and interventions that can alter the number of contacts and infectivity in the setting being modelled.

We assumed that all susceptible persons have an equal probability of contacting infected individuals and acquiring the virus (i.e., homogenous mixing). As the epidemic grows, the number of susceptible persons declines. Thus, not all the daily contacts of infected individuals will be with susceptible persons. The daily infection rate for a susceptible person is equal to the sum of transmission rates from all infected persons across all infection states divided by the cohort size. This leads to an expected daily number of infections equal to the number of susceptible persons multiplied by the infection rate on that day.

***Testing and Interventions***

In this analysis, testing is performed on a nasopharyngeal specimen by polymerase chain reaction (PCR) assay. We assumed that test characteristics including sensitivity and specificity are independent of disease state – i.e., the sensitivity is the same for those in the mild/moderate disease state and those in the critical disease state. We assumed that, after providing a specimen for testing and while awaiting the test result, hospitalised individuals are isolated and non-hospitalised individuals are advised to self-isolate at home. In the model, test results are acted upon (an intervention is started) on the day that the result is delivered.

**S1.2. Model Calibration and Validation**

To calibrate our model output with the COVID-19 epidemic in South Africa, we adjusted the transmission multiplier to generate an effective reproduction number ($R_e$) of 1.5, matching that published by South Africa's National Institute for Communicable Diseases (NICD) based on empirical data collected in the country up to 19 May 2020.[4] We also evaluated alternative epidemic growth scenarios with $R_e$=1.1, $R_e$=1.2, or $R_e$=2.6, reflecting a range of estimates from different periods and regions in the NICD report.

For validation, we assumed a SARS-CoV-2 infection prevalence of 0·1% at model initiation, corresponding to approximately 11,000 cases among the KwaZulu-Natal population of 11 million people. We then looked to KwaZulu-Natal data to determine the date at which there were 1,100 reported (confirmed) cases of SARS-CoV-2 infection, assuming that the true prevalence was 10 times higher than the reported number of cases. Data compiled by the University of Pretoria indicated that this occurred on 6 May 2020.[5] This date would thus correspond to "Day 0" in our model. We then compared cumulative deaths through 30 August in the University of Pretoria database with the corresponding Day 116 cumulative deaths in our model. The database indicated 2,100 COVID-19 deaths during that period, while we estimated 2,806 deaths over 116 days in our model with $R_e$=1.5 and the *HT* strategy. Thus, the model output for COVID-19 deaths was similar to the numbers reported in KwaZulu-Natal, considering likely undercounting of COVID-19-related deaths.[6]

#### S1.3. Input Parameters

***Natural History***

We calculated age-stratified disease path probabilities. We used the proportions of people with COVID-19 who were: (a) asymptomatic,[7,8] (b) admitted to the ICU,[9] (c) hospitalised,[9] and (d) undiagnosed,[10] and the age-stratified proportions of different disease severity states.[11]

We used the following sources to derive the duration of time in each state: presymptomatic infectious time;[11,12] duration of viral shedding based on PCR detectability (WHO-China CDC Report,[11] Hu et al.,[13] Zhou et al.[14]); time to development of pneumonia (Wang et al.[15]); time to ICU admission (Zhou et al.[14]); time spent in the ICU (Zhou et al.[14]); and median time to death (Zhou et al.[14]). We calculated the transition probabilities until recovery (defined as the end of viral shedding) and the transition probabilities between disease states including death. Subsequently, after determining the duration in each state, we estimated transition rates. We then calculated transition probabilities from the transition rates.

$$Transition\ rate = rt = \frac{1}{duration\ of\ the\ transition}.$$

$$Transition\ probability = p = 1 - \exp(-rt).$$

***Life Expectancy and Years-of-Life Lost***

We estimated the years-of-life saved (YLS) from each averted death from COVID-19 in KwaZulu-Natal, South Africa. To do this, we calculated years-of-life lost (YLL), defined as the average number of years a person would have lived had s/he not died from COVID-19.[16] The absolute number of YLL were:[16,17]

$$YLL_{age\ i} = Deaths_{age\ i} * LE_{age\ i}$$

Where,
$Deaths_{age\ i}$ is the number of deaths from COVID-19 in the age stratum,
$LE_{age\ i}$ is the life expectancy in South Africa in the age stratum.

Therefore, age-stratified deaths and age-stratified life expectancy are needed. We obtained or calculated these data from the following sources,

1. Age-stratified distribution of cases: We used the published South Africa National Institute for Communicable Disease for Communicable Diseases (NICD) COVID-19 epidemiology report.[18]

2. Age-stratified distribution of deaths: We used data from the South Africa NICD COVID-19 update report.[19]

3. Calculate life expectancy: Published South Africa life tables are stratified by sex. Our model analysis was not stratified by sex. Therefore, we generated a standard abridged life table, not stratified by sex.
   I. To create a life table for South Africa, we used the following data:
      a. All-cause mortality: World Health Organization disease burden and mortality[20]
      b. Age- and sex-stratified population size: United Nations World Population Prospects 2019[21]
   II. Using SAS software (Cary, North Carolina, USA), we generated a life table. From this, we estimated the expected life-years at any given age.

4. Calculate the age-stratified absolute number of YLL:

$$YLL_{age\ i} = Deaths_{age\ i} * LE_{age\ i}$$

5. Calculate the total absolute number of YLL, base case:

$$YLL_{base\ case} = \sum YLL_{age\ i}$$

6. Calculate the mean YLL:

$$Mean\ YLL = \frac{\sum YLL_{age\ i}}{\sum Deaths_{age\ i}}$$

7. Calculate the absolute number of YLL associated with different intervention strategies: We used the mean YLL to estimate intervention-specific YLL

$$YLL_{intervention\ j} = Mean\ YLL * Deaths_{intervention\ j}$$

The estimates for YLL for each COVID-19 death were 16.8 (undiscounted) and 12.5 (discounted 3%/year).

***Transmission***
Assuming that (a) $R_0$ is 2.6 for individuals with asymptomatic and mild/moderate disease, (b) $R_0$ is one-tenth of 2.6 for individuals with severe and critical disease,[22] and (c) viral shedding times are 9.5, 12, 19, and 24 days for individuals with asymptomatic, mild/moderate, severe, and critical disease, respectively,[11,13,14] we estimated the nominal transmission rate as described above in S1.1.

***Resource Utilization and Costs***
We applied costs from the health sector perspective. We adjusted costs to 2019 United States dollars, using South Africa-specific inflation and exchange rates.[2,3] We obtained costs of clinical care from Mahomed et al. and Netcare Hospitals.[23,24] We obtained the cost of PCR testing, including personnel and supplies, from the Africa Health Research Institute (personal communication). Costs and sources are indicated in Tables S12-S16.

We derived the number of ICU and non-ICU hospital beds available in KwaZulu-Natal (KZN) based on data reported by the South Africa Department of Health:

$$(a)\quad ICU\ hospital\ beds_{KZN} = \frac{Total\ (non-ICU\ and\ ICU)\ hospital\ beds_{KZN}}{Total\ (non-ICU\ and\ ICU)\ hospital\ beds_{South\ Africa}} \times ICU\ hospital\ beds_{South\ Africa}$$

$$(b)\quad non-ICU\ hospital\ beds_{KZN} = \frac{Total\ (non-ICU\ and\ ICU)\ hospital\ beds_{KZN}}{Total\ (non-ICU\ and\ ICU)\ hospital\ beds_{South\ Africa}} \times non-ICU\ hospital\ beds_{South\ Africa}$$

We derived the costs of additional intervention strategies from data supplied by the Africa Health Research Institute. The daily per-person costs of isolation and quarantine centre beds were based on the cost of a 500-person tent and personnel requirements.

To calculate the per-person cost of contact tracing and mass symptom screening, including personnel, supplies, and transportation, we assumed that community health workers could visit 30 households per day, with 5 individuals per house, on 20 days per month:

$$Per\ person\ contact\ tracing\ cost = \frac{Monthly\ cost\ of\ contact\ tracing}{Days\ per\ month\ \times Households\ per\ day\ \times\ Individuals\ per\ house}$$

(the same per-person cost was applied for mass symptom screening)

We assumed that the per-unit costs of resources would be the same regardless of the total quantity. For example, per-test cost of performing a PCR assay was the same regardless of the number of PCR assays performed, and per-person daily cost of a stay at an isolation centre was the same regardless of the number of individuals housed at an isolation centre.

Costs of the various interventions included expenses associated with personnel, supplies, personal protective equipment, and transportation of specimens and personnel. We did not account for additional costs of staff training. The per-test cost of a PCR assay included the cost of reagents and personnel and specimen transportation, but not the cost of additional machines or training new technicians. To reflect uncertainty in our estimates, we varied costs between 50% and 200% of their base case value in sensitivity analyses.

***Mass Symptom Screening Efficacy***

To calculate the increase in the cumulative probability of undergoing testing from mass symptom screening (MS) relative to contact tracing (CT), we assumed that MS would screen the population of 11 million twice per year. We assumed that individuals with mild/moderate symptoms are symptomatic for 10 days, on average:

$$Increase\ in\ MS\ efficacy\ relative\ to\ CT = \frac{Average\ duration\ of mild/moderate\ symptoms\ \times screens\ per\ year}{Time\ (days)}$$

$$Increase\ in\ MS\ efficacy\ relative\ to\ CT = \frac{10\ \times\ 2}{360} = 5.6\%$$

***Influenza-like Illness in Mass Symptom Screening***

Based on a cross-sectional household survey conducted in KwaZulu-Natal by the Africa Health Research Institute, approximately 1% of individuals have symptoms of an influenza-like illness (ILI).[26] To calculate the number of individuals with ILI who would be tested under MS each day:

$$Individuals\ with\ ILI\ tested\ under\ MS, daily = \frac{Individuals\ screened\ under\ MS\ \times screens\ per\ year}{Time\ (days)} \times\ prevalence\ of\ ILI$$

$$Individuals\ with\ ILI\ tested\ under\ MS, daily = \frac{11{,}000{,}000 \times 2}{360} \times\ 1\% = 611$$

***Tracing of Non-infected Contacts***

The number of non-infected individuals who present to care due to contact tracing is linked to the number of positive PCR tests on a given day in our model – the event that initiates a contact trace. The 26 July 2020 COVID-19 KwaZulu-Natal situation report contained the following data related to contact tracing:[27]

| Description | Value |
|---|---|
| Total cases | 64,061 |
| Number of contacts identified, traced, and tested | 50,757 |
| Number of contacts testing positive | 2,152 |
| Number of contacts testing negative | 48,605 |

We assumed that a PCR test has a sensitivity of 70% and specificity of 100%, which implies that of 50,757 contacts identified, traced, and tested, approximately 3,074 are infected with SARS-CoV-2 and 47,683 are not. We then derived the expected number of non-infected contacts traced per positive PCR result ($\eta_0$) by dividing the number of non-infected contacts traced by the number of "original" confirmed cases of COVID-19 in the 26 July 2020 situation report.[27]

$$\eta_0 = \frac{47{,}683}{64{,}061 - 2{,}152} = 0.77$$

We modified the expected number of non-infected contacts traced per positive PCR result to reflect the prevalence of active disease within the population as follows:

$$\eta(t) = \eta_0 \cdot \frac{S(t) + R(t)}{N_0}$$

where $S(t)$ and $R(t)$ represent the number of susceptible and recovered individuals on day $t$, and $N_0$ represents the population of KwaZulu-Natal. The number of non-infected contacts traced on day $t$ is given by

$$N_{CT}(t) = \eta(t) \cdot N_{PCR}^{+}(t - \theta)$$

where $N_{PCR}^{+}(t-\theta)$ represents the number of positive PCR tests on day $t-\ \theta$, where $\theta$ represents the number of days it takes to trace an individual's contacts (assumed to be two days). Once traced, non-infected contacts may incur costs related to PCR testing and quarantine centres in the same manner as infected individuals.

#### APPENDIX REFERENCES

1 Edoka IP, Stacey NK. Estimating a cost-effectiveness threshold for health care decision-making in South Africa. *Health Policy Plan* 2020; **35**: 546–55.

2 Board of Governors of the Federal Reserve System (US). South Africa / U.S. Foreign Exchange Rate. FRED, Federal Reserve Bank of St. Louis. https://fred.stlouisfed.org/series/DEXSFUS (accessed Sept 8, 2020).

3 Organization for Economic Co-operation and Development. Consumer Price Index: All Items for South Africa. FRED, Federal Reserve Bank of St. Louis. https://fred.stlouisfed.org/series/ZAFCPIALLMINMEI (accessed Sept 8, 2020).

4 National Institute for Communicable Diseases. The Initial and Daily COVID-19 Effective Reproductive Number (R) in South Africa. 2020; published online May 27. https://www.nicd.ac.za/wp-content/uploads/2020/05/The-Initial-and-Daily-COVID-19-Effective-Reproductive-Number-R-in-South-Africa-002.pdf (accessed June 9, 2020).

5 Data Science for Social Impact Research Group @ University of Pretoria. Coronavirus COVID-19 (2019-nCoV) Data Repository and Dashboard for South Africa. 2020; published online Sept 8. https://github.com/dsfsi/covid19za (accessed Sept 8, 2020).

6 South African Medical Research Council. Report on Weekly Deaths in South Africa. South African Medical Research Council. 2020; published online Aug 26. https://www.samrc.ac.za/reports/report-weekly-deaths-south-africa (accessed Sept 8, 2020).

7 Mizumoto K, Kagaya K, Zarebski A, Chowell G. Estimating the asymptomatic proportion of coronavirus disease 2019 (COVID-19) cases on board the Diamond Princess cruise ship, Yokohama, Japan, 2020. *Eurosurveillance* 2020; **25**. DOI:10.2807/1560-7917.ES.2020.25.10.2000180.

8 Tao Y, Cheng P, Chen W, *et al.* High incidence of asymptomatic SARS-CoV-2 infection, Chongqing, China. Emergency Medicine, 2020 DOI:10.1101/2020.03.16.20037259.

9 Massachusetts Department of Health. COVID-19 Dashboard. 2020. https://www.mass.gov/doc/covid-19-dashboard-april-20-2020/download (accessed June 10, 2020).

10 Rothwell J. Estimating COVID-19 Prevalence in Symptomatic Americans. 2020; published online April 3. https://news.gallup.com/opinion/gallup/306458/estimating-covid-prevalence-symptomatic-americans.aspx (accessed June 10, 2020).

11 WHO-China Joint Mission on Coronavirus Diseaes 2019 (COVID-19). Report of the WHO-China Joint Mission on Coronavirus Diseases 2019 (COVID-19). 2020. https://www.who.int/docs/default-source/coronaviruse/who-china-joint-mission-on-covid-19-final-report.pdf (accessed May 27, 2020).

12 He X, Lau EHY, Wu P, *et al.* Temporal dynamics in viral shedding and transmissibility of COVID-19. *Nat Med* 2020; **26**: 672–5.

13 Hu Z, Song C, Xu C, *et al.* Clinical characteristics of 24 asymptomatic infections with COVID-19 screened among close contacts in Nanjing, China. *Sci China Life Sci* 2020; **63**: 706–11.

14 Zhou F, Yu T, Du R, *et al.* Clinical course and risk factors for mortality of adult inpatients with COVID-19 in Wuhan, China: a retrospective cohort study. *Lancet* 2020; **395**: 1054–62.

15 Wang D, Hu B, Hu C, *et al.* Clinical characteristics of 138 hospitalized patients with 2019 novel coronavirus–infected pneumonia in Wuhan, China. *JAMA* 2020; **323**: 1061.

16 Gardner JW, Sanborn JS. Years of potential life lost (YPLL)--what does it measure? *Epidemiology* 1990; **1**: 322–9.

17 Martinez R, Soliz P, Caixeta R, Ordunez P. Reflection on modern methods: years of life lost due to premature mortality-a versatile and comprehensive measure for monitoring non-communicable disease mortality. *Int J Epidemiol* 2019; **48**: 1367–76.

18 National Institute for Communicable Diseases, South Africa. COVID-19 Weekly Epidemiology Brief. https://www.nicd.ac.za/wp-content/uploads/2020/05/Week-18-Weekly-Epidemiology-Brief-Template-V8.pdf (accessed Sept 3, 2020).

19 National Institute for Communicable Diseases, South Africa. COVID-19 Update. NICD. 2020; published online May 2. https://www.nicd.ac.za/covid-19-update-46/ (accessed Sept 3, 2020).

20 World Health Organization. Disease Burden and Mortality Estimates: Cause-Specific Mortality, 2000-2016. 2017. https://www.nicd.ac.za/covid-19-update-36/ (accessed June 10, 2020).

21 United Nations. World Population Prospects, South Africa. 2019. https://population.un.org/wpp/Download/Standard/Population/ (accessed June 10, 2020).

22 Liu Y, Gayle AA, Wilder-Smith A, Rocklöv J. The reproductive number of COVID-19 is higher compared to SARS coronavirus. *J Travel Med* 2020; **27**. DOI:10.1093/jtm/taaa021.

23 Mahomed S, Mahomed OH. Cost of intensive care services at a central hospital in South Africa. *S Afr Med J* 2018; **109**: 35.

24 Netcare Hospitals. Netcare Tariffs. 2016. https://www.netcarehospitals.co.za/ (accessed June 10, 2020).

25 National Department of Health, South Africa. COVID-19 Public Health Response. 2020; published online April 10. https://sacoronavirus.co.za/2020/04/11/covid-19-public-health-response/ (accessed June 10, 2020).

26 Siedner MJ, Harling G, Derache A, *et al.* Protocol: Leveraging a demographic and health surveillance system for Covid-19 Surveillance in rural KwaZulu-Natal. *Wellcome Open Res* 2020; **5**: 109.

27 Ponch Bapela M, Mhlongo B, editors. KwaZulu-Natal Department of Health COVID-19 Situational Report: 26 July 2020. .

28 World Health Organization. Choosing interventions that are cost effective (WHO-CHOICE). http://www.who.int/choice/en/ (accessed April 28, 2020).

29 Reddy KP, Gupta-Wright A, Fielding KL, *et al.* Cost-effectiveness of urine-based tuberculosis screening in hospitalised patients with HIV in Africa: a microsimulation modelling study. *Lancet Glob Health* 2019; **7**: e200–8.

30 Craig J, Kalanxhi E, Hauck S. National estimates of critical care capacity in 54 African countries. Public and Global Health, 2020 DOI:10.1101/2020.05.13.20100727.

**Table S1. Additional natural history input parameters for a model-based analysis of COVID-19 intervention strategies in KwaZulu-Natal, South Africa.**

| Parameter | Value | | | | Source |
|---|---|---|---|---|---|
| Disease path probability*, stratified by age, % | Asymp. | Mild/Mod. | Severe | Critical | † |
| 0-19y | 29.93 | 69.78 | 0.25 | 0.03 | |
| 20-59y | 17.90 | 80.38 | 0·80 | 0.93 | |
| ≥60y | 17.10 | 76.37 | 1.40 | 5.16 | |
| Duration of health states, stratified by disease path, days | Asymp. | Mild/Mod. | Severe | Critical | † |
| Pre-infectious latency | 2.6 | 2.6 | 2.6 | 2.6 | |
| Asymptomatic | 9.5 | 2.0 | 2.0 | 2.0 | |
| Mild/moderate disease | -- | 10.0 | 6.5 | 3.0 | |
| Severe disease | -- | -- | 10.5 | 7.1 | |
| Critical disease | -- | -- | -- | 11.9 | |
| Recuperation after critical disease | -- | -- | -- | 5.7 | |
| Mortality probability among those with critical COVID-19 disease, stratified by age, daily, % | 0-19y | 20-59y | ≥60y | | † |
| Without hospital care | 11.7500 | 16.6200 | 20.3300 | | |
| With hospital care | 0.0006 | 0.3800 | 5.0000 | | |

COVID-19: coronavirus disease 2019. y: years. Asymp.: asymptomatic. Mod.: moderate.

*Disease path probability refers to the likelihood that an individual, once infected with SARS-CoV-2, will eventually progress to the specified COVID-19 disease state.

†Derivation of natural history parameters is described in the appendix, p.5.

**Table S2. Intervention-related input parameters for a model-based analysis of COVID-19 intervention strategies in KwaZulu-Natal, South Africa.**

| Intervention Strategies | *HT* | *HT+CT* | *HT+CT +IC* | *HT+CT +IC+MS* | *HT+CT +IC+QC* | *HT+CT +IC+MS+QC* | Source |
|---|---|---|---|---|---|---|---|
| Cumulative probability of undergoing testing, over health state duration, % | | | | | | | |
| Susceptible | 0 | Variable | Variable | Variable | Variable | Variable | * |
| Pre-infectious latency | 0 | 10 (5-20) | 10 (5-20) | 12.5 (6.25-25) | 10 (5-20) | 12.5 (6.25-25) | Asm. |
| Asymptomatic | 0 | 10 (5-20) | 10 (5-20) | 12.5 (6.25-25) | 10 (5-20) | 12.5 (6.25-25) | Asm. |
| Mild/moderate disease | 30 | 35 (33-40) | 35 (33-40) | 40 (35-50) | 35 (33-40) | 40 (35-50) | Asm. |
| Severe disease | 100 | 100 | 100 | 100 | 100 | 100 | Asm. |
| Critical disease | 100 | 100 | 100 | 100 | 100 | 100 | Asm. |
| Recovered | 0 | Variable | Variable | Variable | Variable | Variable | * |
| Reduction in onward transmission, % (range) | | | | | | | |
| Home isolation/quarantine | 50 (25-75) | 50 (25-75) | 50 (25-75) | 50 (25-75) | -- | -- | Asm. |
| Isolation centre | -- | -- | 95 (75-99) | 95 (75-99) | 95 (75-99) | 95 (75-99) | Asm. |
| Quarantine centre | -- | -- | -- | -- | 95 (75-99) | 95 (75-99) | Asm. |

COVID-19: coronavirus disease 2019. Asm.: assumption. HT: healthcare testing. CT: contact tracing within households. IC: isolation centre. MS: mass symptom screen. QC: quarantine centre.

Values indicated are those applied in the base case analyses or, in parentheses, the ranges evaluated in sensitivity analysis.

*Testing among those in the susceptible or recovered states is described in the appendix, p.7-8.

**Table S3. Sensitivity analysis: varying the costs of contact tracing and mass symptom screen strategies.**

| Cost | Strategy | Total life-years lost, n | Total health care costs over 360 days, 2019 USD | ICER, 2019 USD/YLS |
|---|---|---|---|---|
| Base case | *HT* | 450,940 | 437,000,000 | -- |
| | *HT+CT+IC+MS+QC* | 27,220 | 581,000,000 | 340 |
| | *HT+CT* | 322,970 | 588,000,000 | DOMINATED |
| | *HT+CT+IC+MS* | 60,930 | 668,000,000 | DOMINATED |
| | *HT+CT+IC* | 128,890 | 780,000,000 | DOMINATED |
| | *HT+CT+IC+QC* | 60,190 | 965,000,000 | DOMINATED |
| Contact tracing and mass symptom screening cost changed to 50% of base case value | *HT* | 450,940 | 437,000,000 | -- |
| | *HT+CT+IC+MS+QC* | 27,220 | 551,000,000 | 270 |
| | *HT+CT* | 322,970 | 584,000,000 | DOMINATED |
| | *HT+CT+IC+MS* | 60,930 | 637,000,000 | DOMINATED |
| | *HT+CT+IC* | 128,890 | 778,000,000 | DOMINATED |
| | *HT+CT+IC+QC* | 60,190 | 963,000,000 | DOMINATED |
| Contact tracing and mass symptom screening cost changed to 200% of base case value | *HT* | 450,940 | 437,000,000 | -- |
| | *HT+CT* | 322,970 | 596,000,000 | dominated |
| | *HT+CT+IC+MS+QC* | 27,220 | 640,000,000 | 480 |
| | *HT+CT+IC+MS* | 60,930 | 729,000,000 | DOMINATED |
| | *HT+CT+IC* | 128,890 | 786,000,000 | DOMINATED |
| | *HT+CT+IC+QC* | 60,190 | 970,000,000 | DOMINATED |

USD: United States dollars. ICER: incremental cost-effectiveness ratio. YLS: year-of-life saved. HT: healthcare testing. CT: contact tracing within households. IC: isolation centre. MS: mass symptom screen. QC: quarantine centre. DOMINATED: strong dominance, resulting in more life-years lost and higher costs than an alternative strategy. dominated: extended dominance, resulting in an ICER higher than that of an alternative strategy that results in fewer life-years lost.

The ICER is the difference between two strategies in costs divided by the difference in life-years. The displayed life-years and costs are rounded, but the ICER was calculated with non-rounded life-years and costs.

Strategies are listed in order of ascending costs, per convention of cost-effectiveness analysis.

In the base case, contact tracing and mass symptom screening cost $3/person.

**Table S4. Sensitivity analysis: varying the cost of hospitalisation.**

| Cost | Strategy | Total life-years lost, n | Total health care costs over 360 days, 2019 USD | ICER, 2019 USD/YLS |
|---|---|---|---|---|
| Base case | *HT* | 450,940 | 437,000,000 | -- |
| | *HT+CT+IC+MS+QC* | 27,220 | 581,000,000 | 340 |
| | *HT+CT* | 322,970 | 588,000,000 | DOMINATED |
| | *HT+CT+IC+MS* | 60,930 | 668,000,000 | DOMINATED |
| | *HT+CT+IC* | 128,890 | 780,000,000 | DOMINATED |
| | *HT+CT+IC+QC* | 60,190 | 965,000,000 | DOMINATED |
| Hospital (non-IC) bed daily cost changed to WHO estimate ($56/day)* | *HT* | 450,940 | 381,000,000 | -- |
| | *HT+CT* | 322,970 | 535,000,000 | dominated |
| | *HT+CT+IC+MS+QC* | 27,220 | 568,000,000 | 440 |
| | *HT+CT+IC+MS* | 60,930 | 641,000,000 | DOMINATED |
| | *HT+CT+IC* | 128,890 | 743,000,000 | DOMINATED |
| | *HT+CT+IC+QC* | 60,190 | 938,000,000 | DOMINATED |
| Hospital (non-ICU) bed daily cost changed to 50% of base case value | *HT* | 450,940 | 395,000,000 | -- |
| | *HT+CT* | 322,970 | 548,000,000 | dominated |
| | *HT+CT+IC+MS+QC* | 27,220 | 571,000,000 | 420 |
| | *HT+CT+IC+MS* | 60,930 | 647,000,000 | DOMINATED |
| | *HT+CT+IC* | 128,890 | 752,000,000 | DOMINATED |
| | *HT+CT+IC+QC* | 60,190 | 945,000,000 | DOMINATED |
| Hospital (non-ICU) bed daily cost changed to 200% of base case value | *HT* | 450,940 | 521,000,000 | -- |
| | *HT+CT+IC+MS+QC* | 27,220 | 600,000,000 | 190 |
| | *HT+CT* | 322,970 | 669,000,000 | DOMINATED |
| | *HT+CT+IC+MS* | 60,930 | 709,000,000 | DOMINATED |
| | *HT+CT+IC* | 128,890 | 837,000,000 | DOMINATED |
| | *HT+CT+IC+QC* | 60,190 | 1,007,000,000 | DOMINATED |
| ICU bed daily cost changed to 50% of base case value | *HT* | 450,940 | 281,000,000 | -- |
| | *HT+CT* | 322,970 | 419,000,000 | dominated |
| | *HT+CT+IC+QC+MS* | 27,220 | 521,000,000 | 570 |
| | *HT+CT+IC+MS* | 60,930 | 541,000,000 | DOMINATED |
| | *HT+CT+IC* | 128,890 | 609,000,000 | DOMINATED |
| | *HT+CT+IC+QC* | 60,190 | 831,000,000 | DOMINATED |
| ICU bed daily cost changed to 200% of base case value | *HT+CT+IC+MS+QC* | 27,220 | 700,000,000 | -- |
| | *HT* | 450,940 | 748,000,000 | DOMINATED |
| | *HT+CT+IC+MS* | 60,930 | 922,000,000 | DOMINATED |
| | *HT+CT* | 322,970 | 927,000,000 | DOMINATED |
| | *HT+CT+IC* | 128,890 | 1,124,000,000 | DOMINATED |
| | *HT+CT+IC+QC* | 60,190 | 1,234,000,000 | DOMINATED |

USD: United States dollars. ICER: incremental cost-effectiveness ratio. YLS: year-of-life saved. HT: healthcare testing. CT: contact tracing within households. IC: isolation centre. MS: mass symptom screen. QC: quarantine centre. DOMINATED: strong dominance, resulting in more life-years lost and higher costs than an alternative strategy. dominated: extended dominance, resulting in an ICER higher than that of an alternative strategy that results in fewer life-years lost.

The ICER is the difference between two strategies in costs divided by the difference in life-years. The displayed life-years and costs are rounded, but the ICER was calculated with non-rounded life-years and costs.

Strategies are listed in order of ascending costs, per convention of cost-effectiveness analysis.

In the base case, hospital beds cost $165/person/day and ICU beds cost $2,048/person/day.

*This cost is based on a WHO-CHOICE estimate.[28,29]

**Table S5. Sensitivity analysis: varying PCR testing parameters.**

| PCR testing parameter | Strategy | Total life-years lost, n | Total health care costs over 360 days, 2019 USD | ICER, 2019 USD/YLS |
|---|---|---|---|---|
| Base case | *HT* | 450,940 | 437,000,000 | -- |
| | *HT+CT+IC+MS+QC* | 27,220 | 581,000,000 | 340 |
| | *HT+CT* | 322,970 | 588,000,000 | DOMINATED |
| | *HT+CT+IC+MS* | 60,930 | 668,000,000 | DOMINATED |
| | *HT+CT+IC* | 128,890 | 780,000,000 | DOMINATED |
| | *HT+CT+IC+QC* | 60,190 | 965,000,000 | DOMINATED |
| PCR sensitivity changed to 50% | *HT* | 450,940 | 437,000,000 | -- |
| | *HT+CT* | 322,970 | 581,000,000 | dominated |
| | *HT+CT+IC+MS+QC* | 31,850 | 583,000,000 | 350 |
| | *HT+CT+IC+MS* | 78,520 | 672,000,000 | DOMINATED |
| | *HT+CT+IC* | 152,040 | 717,000,000 | DOMINATED |
| | *HT+CT+IC+QC* | 57,590 | 870,000,000 | DOMINATED |
| PCR sensitivity changed to 90% | *HT* | 450,940 | 437,000,000 | -- |
| | *HT+CT* | 322,970 | 596,000,000 | dominated |
| | *HT+CT+IC+MS* | 51,110 | 613,000,000 | 440 |
| | *HT+CT+IC+MS+QC* | 28,150 | 651,000,000 | 1660 |
| | *HT+CT+IC* | 92,410 | 810,000,000 | DOMINATED |
| | *HT+CT+IC+QC* | 60,000 | 956,000,000 | DOMINATED |
| PCR result return time changed to 1 day | *HT* | 563,720 | 495,000,000 | -- |
| | *HT+CT* | 390,750 | 639,000,000 | dominated |
| | *HT+CT+IC+MS+QC* | 23,520 | 653,000,000 | 290 |
| | *HT+CT+IC+MS* | 102,970 | 963,000,000 | DOMINATED |
| | *HT+CT+IC* | 206,300 | 995,000,000 | DOMINATED |
| | *HT+CT+IC+QC* | 56,850 | 1,146,000,000 | DOMINATED |
| PCR result return time changed to 7 days | *HT* | 401,500 | 405,000,000 | -- |
| | *HT+CT+IC+MS* | 65,190 | 537,000,000 | dominated |
| | *HT+CT+IC+MS+QC* | 29,440 | 541,000,000 | 370 |
| | *HT+CT* | 296,860 | 569,000,000 | DOMINATED |
| | *HT+CT+IC* | 118,520 | 691,000,000 | DOMINATED |
| | *HT+CT+IC+QC* | 70,000 | 874,000,000 | DOMINATED |

USD: United States dollars. ICER: incremental cost-effectiveness ratio. YLS: year-of-life saved. PCR: polymerase chain reaction. HT: healthcare testing. CT: contact tracing within households. IC: isolation centre. MS: mass symptom screen. QC: quarantine centre. DOMINATED: strong dominance, resulting in more life-years lost and higher costs than an alternative strategy. dominated: extended dominance, resulting in an ICER higher than that of an alternative strategy that results in fewer life-years lost.

The ICER is the difference between two strategies in costs divided by the difference in life-years. The displayed life-years and costs are rounded, but the ICER was calculated with non-rounded life-years and costs.

Strategies are listed in order of ascending costs, per convention of cost-effectiveness analysis.

In the base case, the PCR test has a 70% sensitivity and a 5-day result return time.

**Table S6. Sensitivity analysis: varying the cost of the PCR test.**

| Cost | Strategy | Total life-years lost, n | Total health care costs over 360 days, 2019 USD | ICER, 2019 USD/YLS |
|---|---|---|---|---|
| Base case | *HT* | 450,940 | 437,000,000 | -- |
| | *HT+CT+IC+MS+QC* | 27,220 | 581,000,000 | 340 |
| | *HT+CT* | 322,970 | 588,000,000 | DOMINATED |
| | *HT+CT+IC+MS* | 60,930 | 668,000,000 | DOMINATED |
| | *HT+CT+IC* | 128,890 | 780,000,000 | DOMINATED |
| | *HT+CT+IC+QC* | 60,190 | 965,000,000 | DOMINATED |
| PCR test cost changed to 50% of base case value | *HT* | 450,940 | 416,000,000 | -- |
| | *HT+CT* | 322,970 | 508,000,000 | dominated |
| | *HT+CT+IC+MS+QC* | 27,220 | 528,000,000 | 260 |
| | *HT+CT+IC+MS* | 60,930 | 605,000,000 | DOMINATED |
| | *HT+CT+IC* | 128,890 | 714,000,000 | DOMINATED |
| | *HT+CT+IC+QC* | 60,190 | 905,000,000 | DOMINATED |
| PCR test cost changed to 200% of base case value | *HT* | 450,940 | 478,000,000 | -- |
| | *HT+CT+IC+MS+QC* | 27,220 | 686,000,000 | 490 |
| | *HT+CT* | 322,970 | 748,000,000 | DOMINATED |
| | *HT+CT+IC+MS* | 60,930 | 793,000,000 | DOMINATED |
| | *HT+CT+IC* | 128,890 | 912,000,000 | DOMINATED |
| | *HT+CT+IC+QC* | 60,190 | 1,086,000,000 | DOMINATED |

USD: United States dollars. ICER: incremental cost-effectiveness ratio. YLS: year-of-life saved. PCR: polymerase chain reaction. HT: healthcare testing. CT: contact tracing within households. IC: isolation centre. MS: mass symptom screen. QC: quarantine centre. DOMINATED: strong dominance, resulting in more life-years lost and higher costs than an alternative strategy. dominated: extended dominance, resulting in an ICER higher than that of an alternative strategy that results in fewer life-years lost.

The ICER is the difference between two strategies in costs divided by the difference in life-years. The displayed life-years and costs are rounded, but the ICER was calculated with non-rounded life-years and costs.

Strategies are listed in order of ascending costs, per convention of cost-effectiveness analysis.

In the base case, the PCR test cost $26/test.

**Table S7. Sensitivity analysis: varying the availability of hospital beds and ICU beds.**

| Number of hospital and ICU beds | Strategy | Peak daily resource use, n | | Total life-years lost, n | Total health care costs over 360 days, 2019 USD | ICER, 2019 USD/YLS |
|---|---|---|---|---|---|---|
| | | Hospital (non-ICU) beds | ICU beds | | | |
| Base case | *HT* | 4,686 | 748 | 450,940 | 437,000,000 | -- |
| | *HT+CT+IC+MS+QC* | 638 | 341 | 27,220 | 581,000,000 | 340 |
| | *HT+CT* | 3,443 | 748 | 322,970 | 588,000,000 | DOMINATED |
| | *HT+CT+IC+MS* | 1,320 | 715 | 60,930 | 668,000,000 | DOMINATED |
| | *HT+CT+IC* | 1,925 | 748 | 128,890 | 780,000,000 | DOMINATED |
| | *HT+CT+IC+QC* | 1,375 | 737 | 60,190 | 965,000,000 | DOMINATED |
| Number of hospital and ICU beds reduced * | *HT* | 4,466 | 374 | 564,280 | 308,000,000 | -- |
| | *HT+CT* | 3,190 | 374 | 438,160 | 447,000,000 | dominated |
| | *HT+CT+IC+MS+QC* | 638 | 341 | 27,220 | 581,000,000 | 510 |
| | *HT+CT+IC+MS* | 1,210 | 374 | 115,000 | 600,000,000 | DOMINATED |
| | *HT+CT+IC* | 1,782 | 374 | 235,380 | 646,000,000 | DOMINATED |
| | *HT+CT+IC+QC* | 1,199 | 374 | 120,740 | 904,000,000 | DOMINATED |

ICU: intensive care unit. USD: United States dollars. ICER: incremental cost-effectiveness ratio. YLS: year-of-life saved. HT: healthcare testing. CT: contact tracing within households. IC: isolation centre. MS: mass symptom screen. QC: quarantine centre. DOMINATED: strong dominance, resulting in more life-years lost and higher costs than an alternative strategy. dominated: extended dominance, resulting in an ICER higher than that of an alternative strategy that results in fewer life-years lost.

The ICER is the difference between two strategies in costs divided by the difference in life-years. The displayed life-years and costs are rounded, but the ICER was calculated with non-rounded life-years and costs.

Strategies are listed in order of ascending costs, per convention of cost-effectiveness analysis.

In the base case, the numbers of available hospital (non-ICU) beds and ICU beds are 26,220 and 748 per 11 million people, respectively.

*We changed the available number of hospital (non-ICU) and ICU beds to match the reported median numbers across countries in sub-Saharan Africa: 22,275 and 374 per 11 million people, respectively.

**Table S8. Sensitivity analysis: varying the effective reproductive number.**

| Effective reproduction number ($R_e$) | Strategy | Total life-years lost, n | Total health care costs over 360 days, 2019 USD | ICER, 2019 USD/YLS |
|---|---|---|---|---|
| 1.1 | *HT+CT+IC+QC* | 2,590 | 110,000,000 | -- |
| | *HT+CT+IC* | 3,700 | 114,000,000 | DOMINATED |
| | *HT+CT* | 8,330 | 127,000,000 | DOMINATED |
| | *HT+CT+IC+MS* | 2,040 | 167,000,000 | dominated |
| | *HT+CT+IC+MS+QC* | 1,300 | 171,000,000 | 47,410 |
| | *HT* | 37,960 | 182,000,000 | DOMINATED |
| 1.2 | *HT+CT+IC+QC* | 3,890 | 139,000,000 | -- |
| | *HT+CT+IC* | 6,850 | 141,000,000 | DOMINATED |
| | *HT+CT+IC+MS* | 4,260 | 183,000,000 | DOMINATED |
| | *HT+CT+IC+QC+MS* | 2,040 | 190,000,000 | 27,590 |
| | *HT+CT* | 32,040 | 276,000,000 | DOMINATED |
| | *HT* | 97,600 | 393,000,000 | DOMINATED |
| 1.5 | *HT* | 450,940 | 437,000,000 | -- |
| | *HT+CT+IC+MS+QC* | 27,220 | 581,000,000 | 340 |
| | *HT+CT* | 322,970 | 588,000,000 | DOMINATED |
| | *HT+CT+IC+MS* | 60,930 | 668,000,000 | DOMINATED |
| | *HT+CT+IC* | 128,890 | 780,000,000 | DOMINATED |
| | *HT+CT+IC+QC* | 60,190 | 965,000,000 | DOMINATED |
| 2.6 | *HT* | 933,730 | 353,000,000 | -- |
| | *HT+CT* | 890,210 | 532,000,000 | 4,130 |
| | *HT+CT+IC* | 838,360 | 1,170,000,000 | dominated |
| | *HT+CT+IC+MS* | 811,510 | 1,317,000,000 | 9,970 |
| | *HT+CT+IC+QC* | 795,580 | 2,380,000,000 | dominated |
| | *HT+CT+IC+MS+QC* | 758,910 | 2,634,000,000 | 25,040 |

USD: United States dollars. ICER: incremental cost-effectiveness ratio. YLS: year-of-life saved. HT: healthcare testing. CT: contact tracing within households. IC: isolation centre. MS: mass symptom screen. QC: quarantine centre. DOMINATED: strong dominance, resulting in more life-years lost and higher costs than an alternative strategy. dominated: extended dominance, resulting in an ICER higher than that of an alternative strategy that results in fewer life-years lost.

The ICER is the difference between two strategies in costs divided by the difference in life-years. The displayed life-years and costs are rounded, but the ICER was calculated with non-rounded life-years and costs.

Strategies are listed in order of ascending costs, per convention of cost-effectiveness analysis.

**Table S9. Sensitivity analysis: varying the efficacies of contact tracing and mass symptom screening.**

| Efficacies of contact tracing and mass symptom screening for case detection | Strategy | Total life-years lost, n | Total health care costs over 360 days, 2019 USD | ICER, 2019 USD/YLS |
|---|---|---|---|---|
| Changed to 50% of base case value (less efficacious) | *HT* | 450,940 | 437,000,000 | -- |
| | *HT+CT* | 393,350 | 582,000,000 | dominated |
| | *HT+CT+IC* | 269,080 | 849,000,000 | dominated |
| | *HT+CT+IC+MS* | 220,560 | 893,000,000 | 1,980 |
| | *HT+CT+IC+QC* | 215,930 | 1,343,000,000 | dominated |
| | *HT+CT+IC+MS+QC* | 143,520 | 1,350,000,000 | 5,930 |
| Base case | *HT* | 450,940 | 437,000,000 | -- |
| | *HT+CT+IC+MS+QC* | 27,220 | 581,000,000 | 340 |
| | *HT+CT* | 322,970 | 588,000,000 | DOMINATED |
| | *HT+CT+IC+MS* | 60,930 | 668,000,000 | DOMINATED |
| | *HT+CT+IC* | 128,890 | 780,000,000 | DOMINATED |
| | *HT+CT+IC+QC* | 60,190 | 965,000,000 | DOMINATED |
| Changed to 200% of base case value (more efficacious) | *HT+CT+IC+QC* | 6,110 | 164,000,000 | -- |
| | *HT+CT+IC+MS+QC* | 2,220 | 183,000,000 | 4,810 |
| | *HT+CT+IC+MS* | 6,110 | 197,000,000 | DOMINATED |
| | *HT+CT+IC* | 20,190 | 282,000,000 | DOMINATED |
| | *HT* | 450,940 | 437,000,000 | DOMINATED |
| | *HT+CT* | 196,860 | 608,000,000 | DOMINATED |

USD: United States dollars. ICER: incremental cost-effectiveness ratio. YLS: year-of-life saved. HT: healthcare testing. CT: contact tracing within households. IC: isolation centre. MS: mass symptom screen. QC: quarantine centre. DOMINATED: strong dominance, resulting in more life-years lost and higher costs than an alternative strategy. dominated: extended dominance, resulting in an ICER higher than that of an alternative strategy that results in fewer life-years lost.

The ICER is the difference between two strategies in costs divided by the difference in life-years. The displayed life-years and costs are rounded, but the ICER was calculated with non-rounded life-years and costs.

Strategies are listed in order of ascending costs, per convention of cost-effectiveness analysis.

**Table S10. Sensitivity analysis: varying the efficacies of isolation and quarantine centres.**

| Efficacies of isolation and quarantine centres in transmission reduction, % | Strategy | Total life-years lost, n | Total health care costs over 360 days, 2019 USD | ICER, 2019 USD/YLS |
|---|---|---|---|---|
| 75 (less efficacious) | *HT* | 450,940 | 437,000,000 | -- |
| | *HT+CT* | 322,970 | 588,000,000 | 1,180 |
| | *HT+CT+IC* | 217,970 | 894,000,000 | dominated |
| | *HT+CT+IC+MS* | 144,630 | 909,000,000 | 1,800 |
| | *HT+CT+IC+MS+QC* | 107,230 | 1,376,000,000 | 12,490 |
| | *HT+CT+IC+QC* | 192,410 | 1,493,000,000 | DOMINATED |
| 95 (base case) | *HT* | 450,940 | 437,000,000 | -- |
| | *HT+CT+IC+MS+QC* | 27,220 | 581,000,000 | 340 |
| | *HT+CT* | 322,970 | 588,000,000 | DOMINATED |
| | *HT+CT+IC+MS* | 60,930 | 668,000,000 | DOMINATED |
| | *HT+CT+IC* | 128,890 | 780,000,000 | DOMINATED |
| | *HT+CT+IC+QC* | 60,190 | 965,000,000 | DOMINATED |
| 99 (more efficacious) | *HT* | 450,940 | 437,000,000 | -- |
| | *HT+CT+IC+MS+QC* | 19,440 | 437,000,000 | 1 |
| | *HT+CT* | 322,970 | 588,000,000 | DOMINATED |
| | *HT+CT+IC+MS* | 51,300 | 614,000,000 | DOMINATED |
| | *HT+CT+IC* | 115,190 | 751,000,000 | DOMINATED |
| | *HT+CT+IC+QC* | 49,630 | 803,000,000 | DOMINATED |

USD: United States dollars. ICER: incremental cost-effectiveness ratio. YLS: year-of-life saved. HT: healthcare testing. CT: contact tracing within households. IC: isolation centre. MS: mass symptom screen. QC: quarantine centre. DOMINATED: strong dominance, resulting in more life-years lost and higher costs than an alternative strategy. dominated: extended dominance, resulting in an ICER higher than that of an alternative strategy that results in fewer life-years lost.

The ICER is the difference between two strategies in costs divided by the difference in life-years. The displayed life-years and costs are rounded, but the ICER was calculated with non-rounded life-years and costs.

Strategies are listed in order of ascending costs, per convention of cost-effectiveness analysis.

**Table S11. Sensitivity analysis: varying the cost of isolation and quarantine centres.**

| Cost of isolation and quarantine centres | Strategy | Total life-years lost, n | Total health care costs over 360 days, 2019 USD | ICER, 2019 USD/YLS |
|---|---|---|---|---|
| Base case | *HT* | 450,940 | 437,000,000 | -- |
| | *HT+CT+IC+MS+QC* | 27,220 | 581,000,000 | 340 |
| | *HT+CT* | 322,970 | 588,000,000 | DOMINATED |
| | *HT+CT+IC+MS* | 60,930 | 668,000,000 | DOMINATED |
| | *HT+CT+IC* | 128,890 | 780,000,000 | DOMINATED |
| | *HT+CT+IC+QC* | 60,190 | 965,000,000 | DOMINATED |
| Isolation centre and quarantine centre costs changed to 25% of base case values | *HT+CT+IC+MS+QC* | 27,220 | 373,000,000 | -- |
| | *HT* | 450,940 | 437,000,000 | DOMINATED |
| | *HT+CT+IC+MS* | 60,930 | 528,000,000 | DOMINATED |
| | *HT+CT+IC+QC* | 60,190 | 568,000,000 | DOMINATED |
| | *HT+CT* | 322,970 | 588,000,000 | DOMINATED |
| | *HT+CT+IC* | 128,890 | 598,000,000 | DOMINATED |
| Isolation centre and quarantine centre costs changed to 50% of base case values | *HT* | 450,940 | 437,000,000 | -- |
| | *HT+CT+IC+MS+QC* | 27,220 | 442,000,000 | 10 |
| | *HT+CT+IC+MS* | 60,930 | 575,000,000 | DOMINATED |
| | *HT+CT* | 322,970 | 588,000,000 | DOMINATED |
| | *HT+CT+IC* | 128,890 | 659,000,000 | DOMINATED |
| | *HT+CT+IC+QC* | 60,190 | 700,000,000 | DOMINATED |
| Isolation centre and quarantine centre costs changed to 200% of base case values | *HT* | 450,940 | 437,000,000 | -- |
| | *HT+CT* | 322,970 | 588,000,000 | dominated |
| | *HT+CT+IC+MS* | 60,930 | 854,000,000 | dominated |
| | *HT+CT+IC+MS+QC* | 27,220 | 858,000,000 | 990 |
| | *HT+CT+IC* | 128,890 | 1,023,000,000 | DOMINATED |
| | *HT+CT+IC+QC* | 60,190 | 1,495,000,000 | DOMINATED |

USD: United States dollars. ICER: incremental cost-effectiveness ratio. YLS: year-of-life saved. HT: healthcare testing. CT: contact tracing within households. IC: isolation centre. MS: mass symptom screen. QC: quarantine centre. DOMINATED: strong dominance, resulting in more life-years lost and higher costs than an alternative strategy. dominated: extended dominance, resulting in an ICER higher than that of an alternative strategy that results in fewer life-years lost.

The ICER is the difference between two strategies in discounted costs divided by the difference in life-years. The displayed life-years and costs are rounded, but the ICER was calculated with non-rounded life-years and costs.

Strategies are listed in order of ascending costs, per convention of cost-effectiveness analysis.

In the base case, isolation centres cost $44/person/day and quarantine centres cost $37/person/day.

**Table S12. Cost of supplies for isolation centres and quarantine centres.**

| Item | Cost, USD* | Quantity | Sub-total, per month, USD | Vendor information |
|---|---|---|---|---|
| Tent assembly and rental | 41,052.63 | 1 | 6,842.11† | David Pam Jang Traders, Durban, KZN |
| Food (3 precooked meals) | 12.00 | 15,000 | 180,000.00 | Functionfoods, Richards Bay, KZN |
| Computers | 1,373.68 | 20 | 4,578.95† | First Technology, Umhlanga, KZN |
| Monitors | 263.16 | 40 | 1,754.39† | First Technology, Umhlanga, KZN |
| Wireless router | 31.53 | 10 | 52.54† | Makro, Springfield, KZN |
| Portable LED light | 11.53 | 100 | 192.11† | Makro, Springfield, KZN |
| Bed | 172.50 | 500 | 14,375.00† | Kendon Medical Supplies (PTY) LTD, Johannesburg, GP |
| Mattress | 43.58 | 500 | 3,631.58† | Surgical and General Supplies, Durban, KZN |
| Bedding | 12.11 | 500 | 1,008.77† | Kendon Medical Supplies (PTY) LTD, Johannesburg, GP |
| Cots | 68.70 | 100 | 1,144.96† | Kendon Medical Supplies (PTY) LTD, Johannesburg, GP |
| Biohazardous waste bin | 10.00 | 100 | 166.69† | Compass Medical Waste Services, Westville, KZN |
| Biohazardous waste bags | 0.35 | 100 | 5.91† | Compass Medical Waste Services, Westville, KZN |
| Refrigerator | 807.84 | 10 | 1,346.40† | Makro, Springfield, KZN |
| Privacy screens | 106.41 | 500 | 8,867.61† | Kendon Medical Supplies (PTY) LTD, Johannesburg, GP |
| File cabinet | 142.05 | 100 | 2,367.54† | Makro, Springfield, KZN |
| Computer desk | 52.58 | 50 | 438.16† | Makro, Springfield, KZN |
| Whiteboard | 47.32 | 20 | 157.72† | Makro, Springfield, KZN |
| Lock box | 133.76 | 50 | 1,114.69† | Kendon Medical Supplies (PTY) LTD, Johannesburg, GP |
| Tape dispenser | 2.88 | 100 | 288.42 | Makro, Springfield, KZN |
| Tape | 1.04 | 100 | 104.21 | Makro, Springfield, KZN |
| General waste bin | 5.53 | 100 | 552.63 | Makro, Springfield, KZN |
| General waste bags | 2.47 | 100 | 247.11 | Makro, Springfield, KZN |
| Cleaning products | 5.66 | 100 | 566.11 | Makro, Springfield, KZN |
| Fire extinguisher | 29.96 | 50 | 1,498.03 | Fire Check, Durban, KZN |
| Laundry service | 1.23 | 500 | 616.46 | Kingsdale Steam Laundry CC, Durban, KZN |
| Portable toilet | 18.16 | 100 | 1,815.79 | Sanitech, Durban, KZN |
| Wheelchair accessible toilet | 92.61 | 20 | 1,852.11 | Sanitech, Durban, KZN |
| Portable toilet (services) | 21.18 | 120 | 2,542.11 | Sanitech, Durban, KZN |
| Gloves | 0.05 | 90,000 | 4,902.63 | Lasec SA (PTY) LTD, Westville, KZN |
| Disposable gowns | 1.97 | 45,000 | 88,519.74 | Surgical and General Supplies, Durban, KZN |
| Face shields | 1.73 | 45,000 | 77,636.84 | Surgical and General Supplies, Durban, KZN |
| Face masks | 0.79 | 75,000 | 59,013.16 | Surgical and General Supplies, Durban, KZN |
| Microwave oven | 63.11 | 10 | 631.05 | Makro, Springfield, KZN |
| Disposable cups | 1.52 | 900 | 1,371.32 | Makro, Springfield, KZN |
| Disposable plates | 2.10 | 900 | 1,892.37 | Makro, Springfield, KZN |
| Portable sink | 42.37 | 500 | 21,184.21 | Sanitech, Durban, KZN |
| Portable sink (services) | 18.16 | 120 | 2,178.95 | Sanitech, Durban, KZN |
| Biohazard spill kit | 47.82 | 100 | 4,781.58 | SpillTech, Congella, KZN |
| Infrared thermometer | 111.97 | 100 | 11,197.37 | Surgical and General Supplies, Durban, KZN |
| Stethoscopes | 2.42 | 500 | 1,210.53 | Kendon Medical Supplies (PTY) LTD, Johannesburg, GP |
| Toilet and sink deliveries | 120.21 | 10 | 1,202.05 | Sanitech, Durban, KZN |

USD: United States dollars. KZN: KwaZulu-Natal, South Africa. GP: Gauteng, South Africa.
*Cost estimates were obtained in May 2020.
†Cost amortized over six months.

**Table S13. Cost of supplies for contact tracing.**

| Item | Cost, USD* | Quantity | Sub-total, per month, USD | Vendor information |
|---|---|---|---|---|
| Infrared thermometer | 111.97 | 2 | 223.95 | Surgical and General Supplies, Durban, KZN |
| Stethoscopes | 2.42 | 2 | 4.48 | Kendon Medical Supplies (PTY) LTD, Johannesburg, GP |
| Gloves | 0.05 | 200 | 10.89 | Lasec SA (PTY) LTD, Westville, KZN |
| Disposable gowns | 1.97 | 600 | 1,180.26 | Surgical and General Supplies, Durban, KZN |
| Face shields | 1.73 | 600 | 1,035.16 | Surgical and General Supplies, Durban, KZN |
| Face masks | 0.79 | 600 | 472.11 | Surgical and General Supplies, Durban, KZN |

USD: United States dollars. KZN: KwaZulu-Natal, South Africa. GP: Gauteng, South Africa.
*Cost estimates were obtained in May 2020.

**Table S14. Personnel costs.**

| Category | Monthly salary, USD* | Quantity | Sub-total, per month, USD | Source |
|---|---|---|---|---|
| Isolation centres | | | | |
| Nurse (junior professional) | 1,494.84 | 40 | 59,793.68 | Median AHRI position payscale |
| Nurse (senior professional) | 2,111.21 | 8 | 16,889.68 | Median AHRI position payscale |
| Nursing assistant | 916.79 | 40 | 36,671.58 | Median AHRI position payscale |
| Janitorial staff (3 days per week) | 697.26 | 10 | 6,972.63 | Median AHRI position payscale |
| Project manager | 2,661.41 | 1 | 2,661.41 | Median AHRI position payscale |
| Unarmed security guard | 1200.51 | 10 | 12,005.05 | Republic Watch Security, Mtubatuba, KZN |
| Quarantine centres | | | | |
| Nurse (junior professional) | 1,494.84 | 5 | 7,474.21 | Median AHRI position payscale |
| Nurse (senior professional) | 2,111.21 | 2 | 4,222.42 | Median AHRI position payscale |
| Nursing assistant | 916.79 | 10 | 9,167.89 | Median AHRI position payscale |
| Janitorial staff (3 days per week) | 697.26 | 5 | 3,486.32 | Median AHRI position payscale |
| Project manager | 2,661.41 | 1 | 2,661.41 | Median AHRI position payscale |
| Unarmed security guard | 1200.51 | 10 | 12,005.05 | Republic Watch Security, Mtubatuba, KZN |
| Contact tracing and mass screening | | | | |
| Nurse (junior professional) | 1,494.84 | 2 | 2,989.68 | Median AHRI position payscale |

USD: United States dollars. KZN: KwaZulu-Natal, South Africa. AHRI: Africa Health Research Institute
*Cost estimates were obtained in May 2020.

**Table S15. Transportation costs.**

| Category | Descriptor / Unit | Value, USD* | Quantity | Sub-total, per month, USD | Source |
|---|---|---|---|---|---|
| Isolation centres | | | | | |
| Transport for 99 staff members | Cost per kilometre | 26.05 | 200 | 5,210.53 | AHRI commercial quote |
| Quarantine centres | | | | | |
| Transport for 23 staff members | Cost per kilometre | 6.05 | 200 | 1,210.53 | AHRI commercial quote |
| Contact tracing and mass screening | | | | | |
| Transport for 2 staff members | Cost per kilometre | 0.26 | 6000 | 1,578.95 | AHRI commercial quote |
| Cost of leasing additional vehicle | Cost per month | 435.22 | 1 | 435.22 | AHRI commercial quote |

USD: United States dollars. AHRI: Africa Health Research Institute
*Cost estimates were obtained in May 2020.

**Table S16. Per-patient costs of testing and interventions.**

| Category | Daily cost per patient, USD | | | | Source |
|---|---|---|---|---|---|
| | Supplies | Personnel | Transportation | Total | |
| Isolation centres | 34.26 | 9.00 | 0.35 | 43.60 | * |
| Quarantine centres | 34.26 | 2.60 | 0.08 | 36.94 | * |
| Contact tracing and mass screening | 0.98 | 1.00 | 0.67 | 2.64 | † |
| Hospital care (non-ICU) | 73.70 | 91.70 | -- | 165.40 | Netcare Hospitals[24] |
| ICU care | 875.00 | 1,089.00 | -- | 1,964.00 | Mahomed et al.[23] |
| Ventilator, mechanical | 93.60 | -- | -- | 93.60 | Netcare Hospitals[24] |
| PCR testing | 26.40 | 0.50 | -- | 26.90 | AHRI communication |

USD: United States dollars. ICU: intensive care unit. PCR: polymerase chain reaction. AHRI: Africa Health Research Institute.

*The per-patient costs of isolation and quarantine centres were estimated based on the total monthly expenses of an alternate care site with the capacity to treat 500 patients daily, for 30 days per month. The total costs included personnel, fixed costs to establish the centres, supplies, and transportation. We assumed that fixed costs were amortized over 6 months (tables S12-S15).

†The per-instance costs of contact tracing and symptom screening were calculated based on the total monthly expenses and screening capacity of a community health worker team (tables S13-S15). We estimated that a two-person team working 20 days per month could conduct approximately 3000 screens per month, visiting an average of 30 five-person households per day.

**Figure S1. Model flowcharts of select COVID-19 epidemic control strategies in KwaZulu-Natal, South Africa.**

(A) *Healthcare Testing*

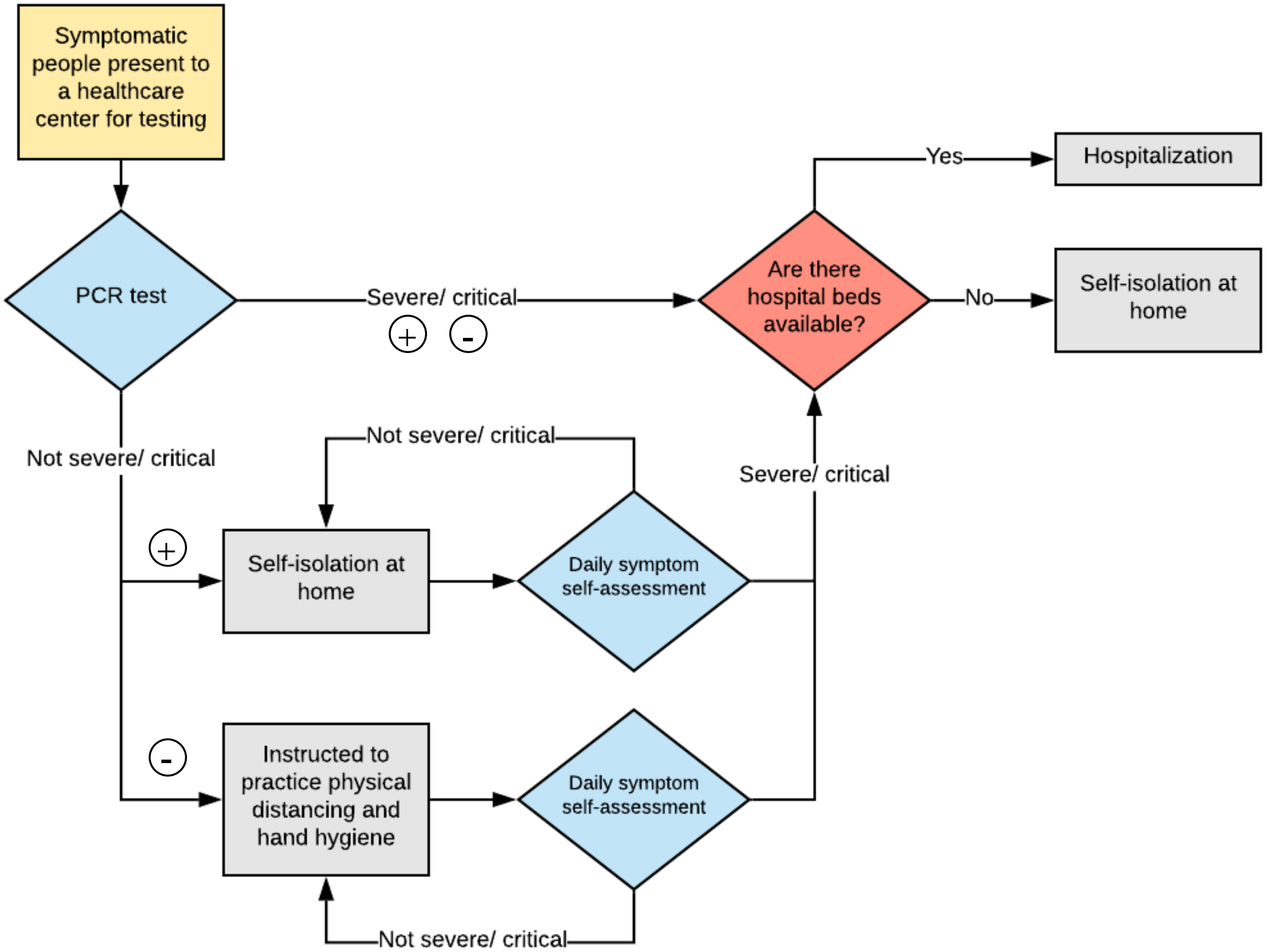


After providing a specimen for testing and while awaiting the result, hospitalised individuals are isolated and non-hospitalised individuals are advised to self-isolate at home. Test results are acted upon (an intervention is started) on the day the result is delivered.

**Figure S1 continued. Model flowcharts of select COVID-19 epidemic control strategies in KwaZulu-Natal, South Africa.**

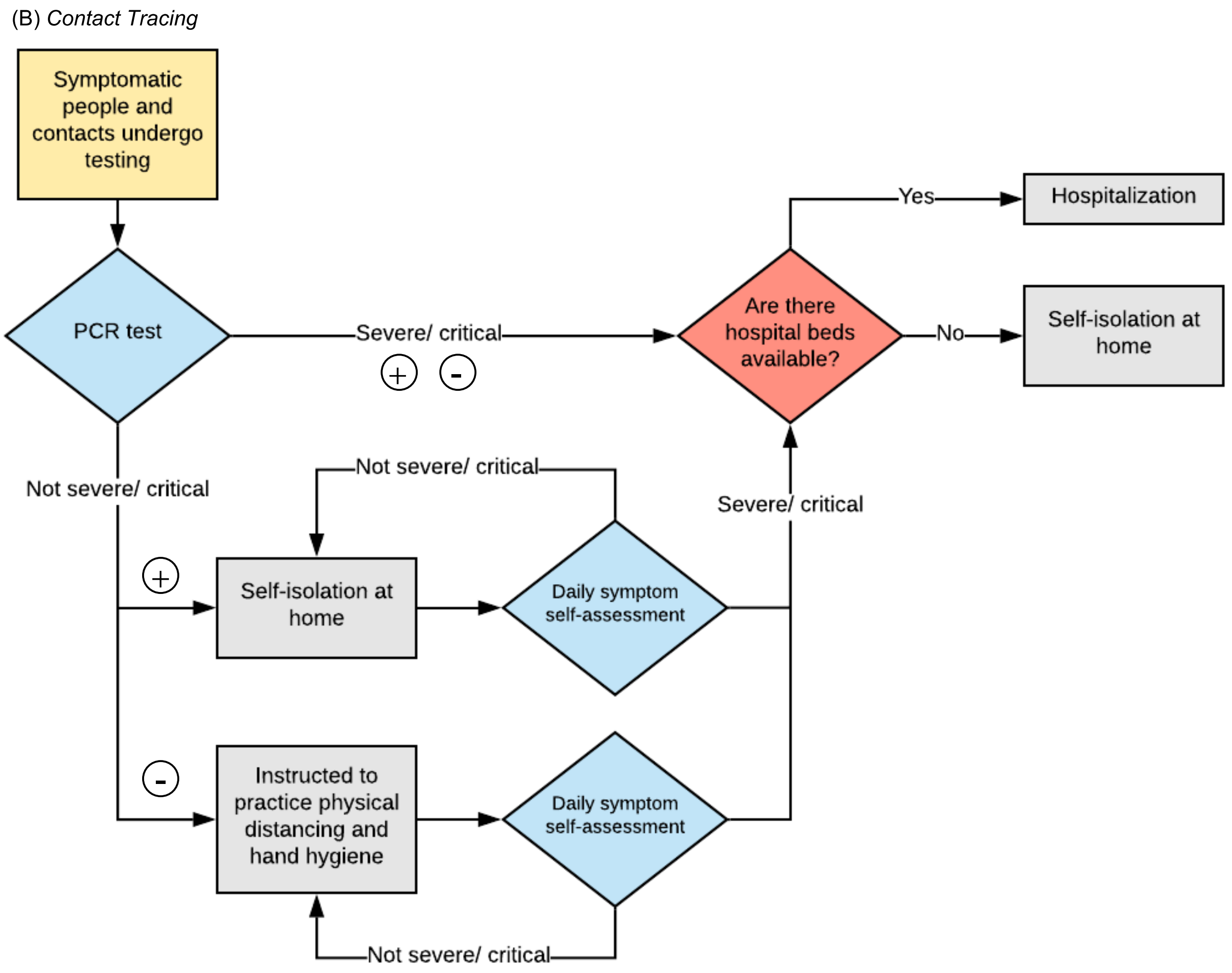


"Contacts" can be symptomatic or asymptomatic. After providing a specimen for testing and while awaiting the result, hospitalised individuals are isolated and non-hospitalised individuals are advised to self-isolate at home. Test results are acted upon (an intervention is started) on the day the result is delivered.

**Figure S1 continued. Model flowcharts of select COVID-19 epidemic control strategies in KwaZulu-Natal, South Africa.**

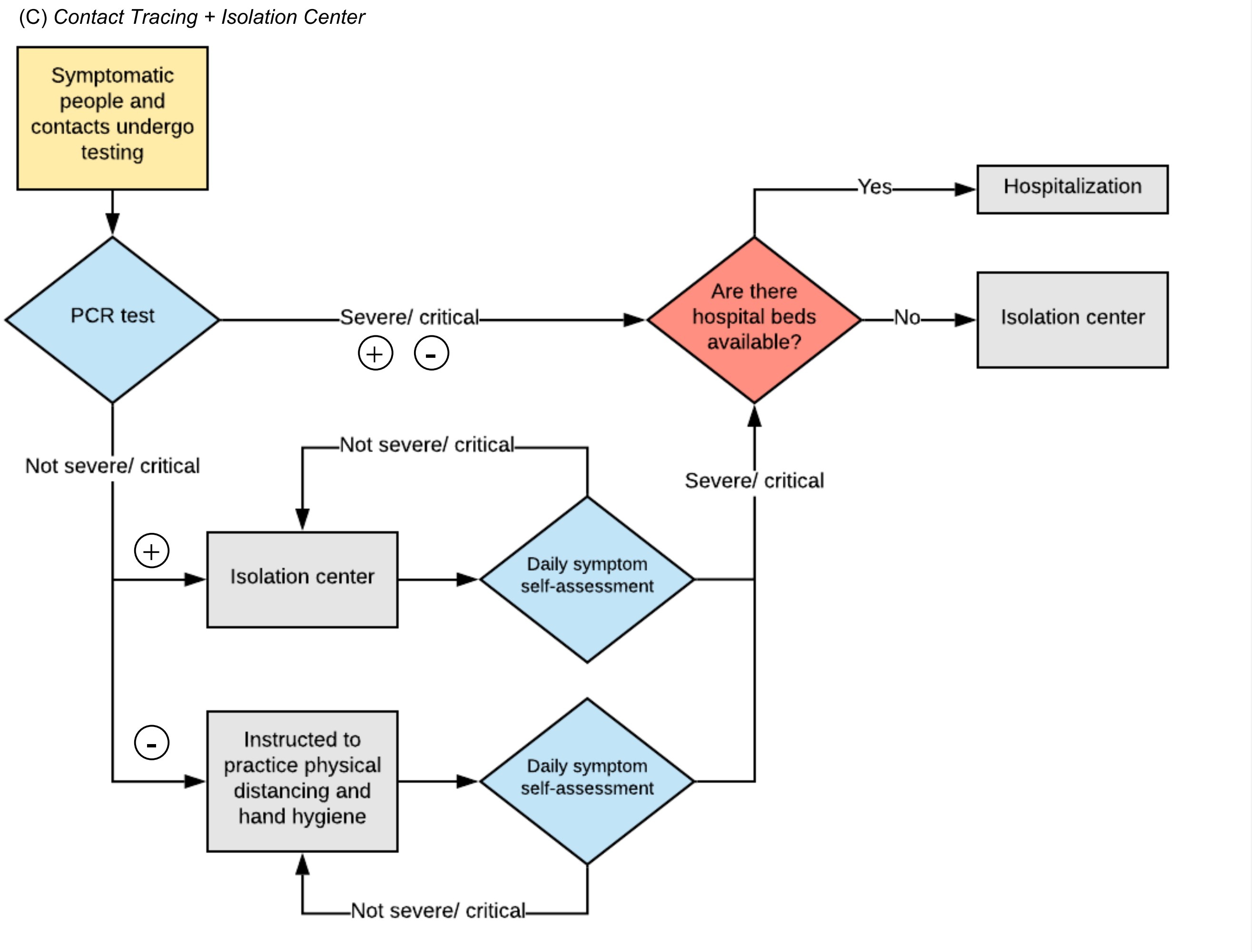


"Contacts" can be symptomatic or asymptomatic. After providing a specimen for testing and while awaiting the result, hospitalised individuals are isolated and non-hospitalised individuals are advised to self-isolate at home. Test results are acted upon (an intervention is started) on the day the result is delivered.

**Figure S1 continued. Model flowcharts of select COVID-19 epidemic control strategies in KwaZulu-Natal, South Africa.**

(D) *Contact Tracing + Isolation Center + Quarantine Center*

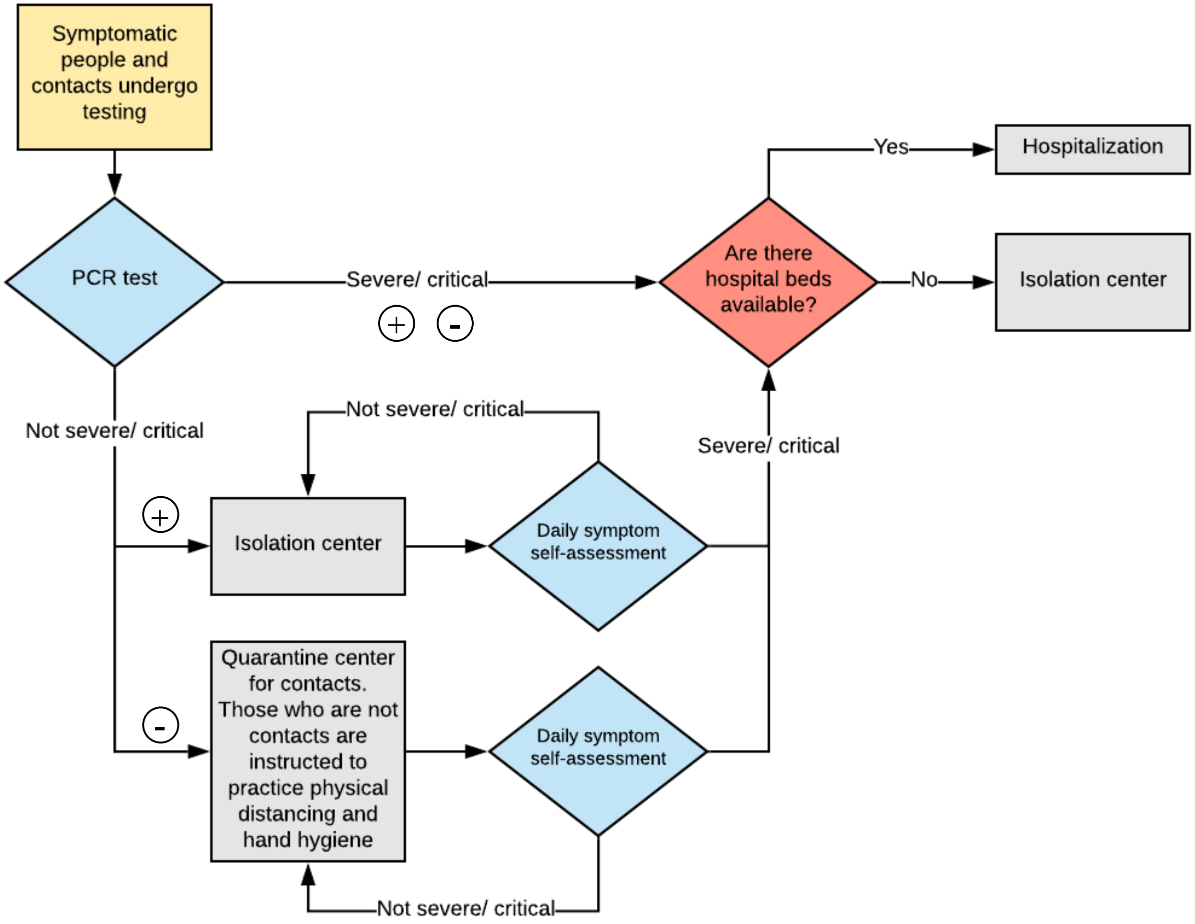


"Contacts" can be symptomatic or asymptomatic. After providing a specimen for testing and while awaiting the result, hospitalised individuals are isolated and non-hospitalised individuals are advised to self-isolate at home. Test results are acted upon (an intervention is started) on the day the result is delivered.

**Figure S2. Illustration of health states and disease paths in the CEACOV model.**

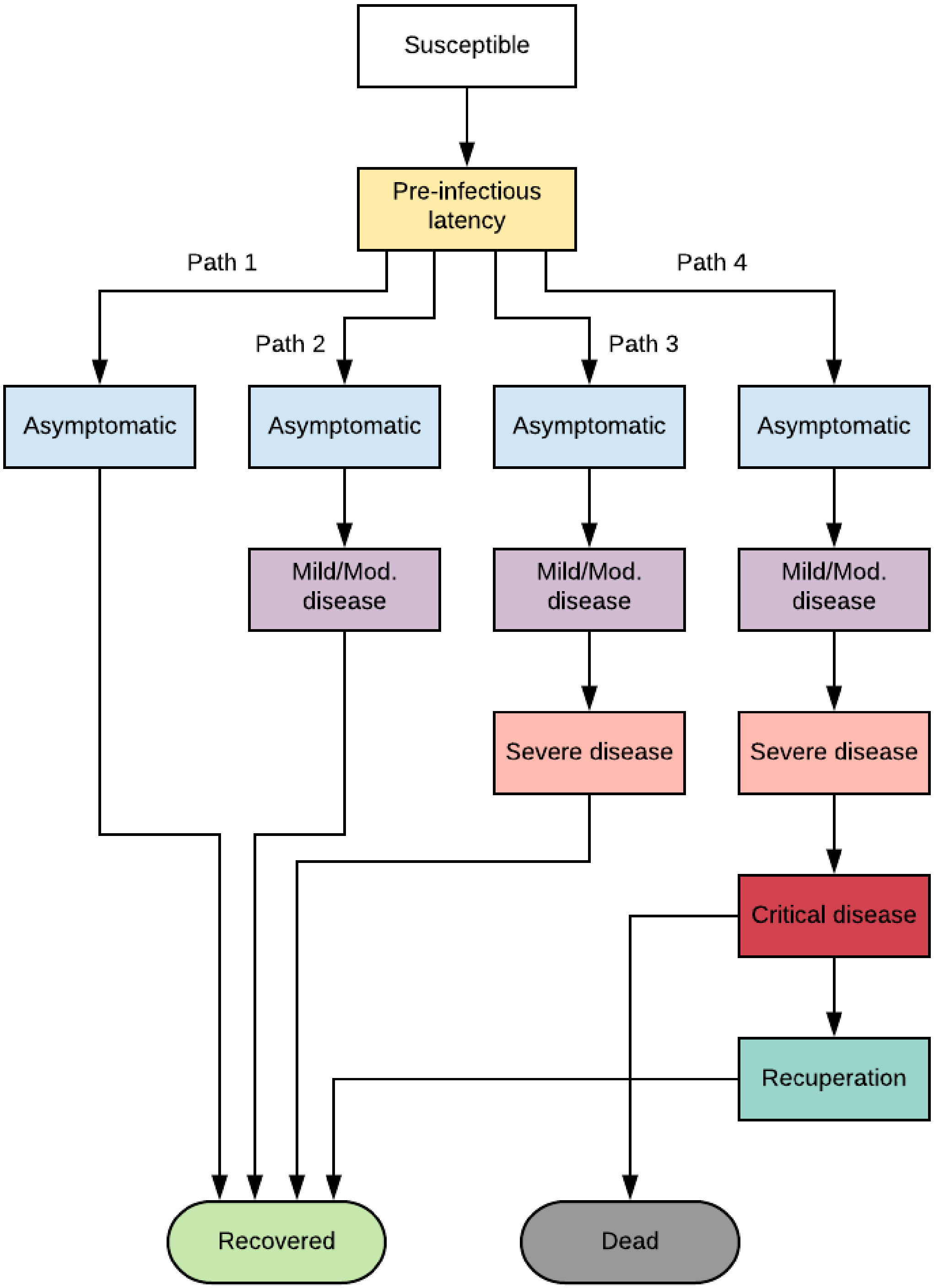


CEACOV: Clinical and Economic Analysis of COVID Interventions. Mod.: moderate.

**Figure S3. Model-projected cumulative and daily SARS-CoV-2 infections by intervention strategy in KwaZulu-Natal, South Africa.**

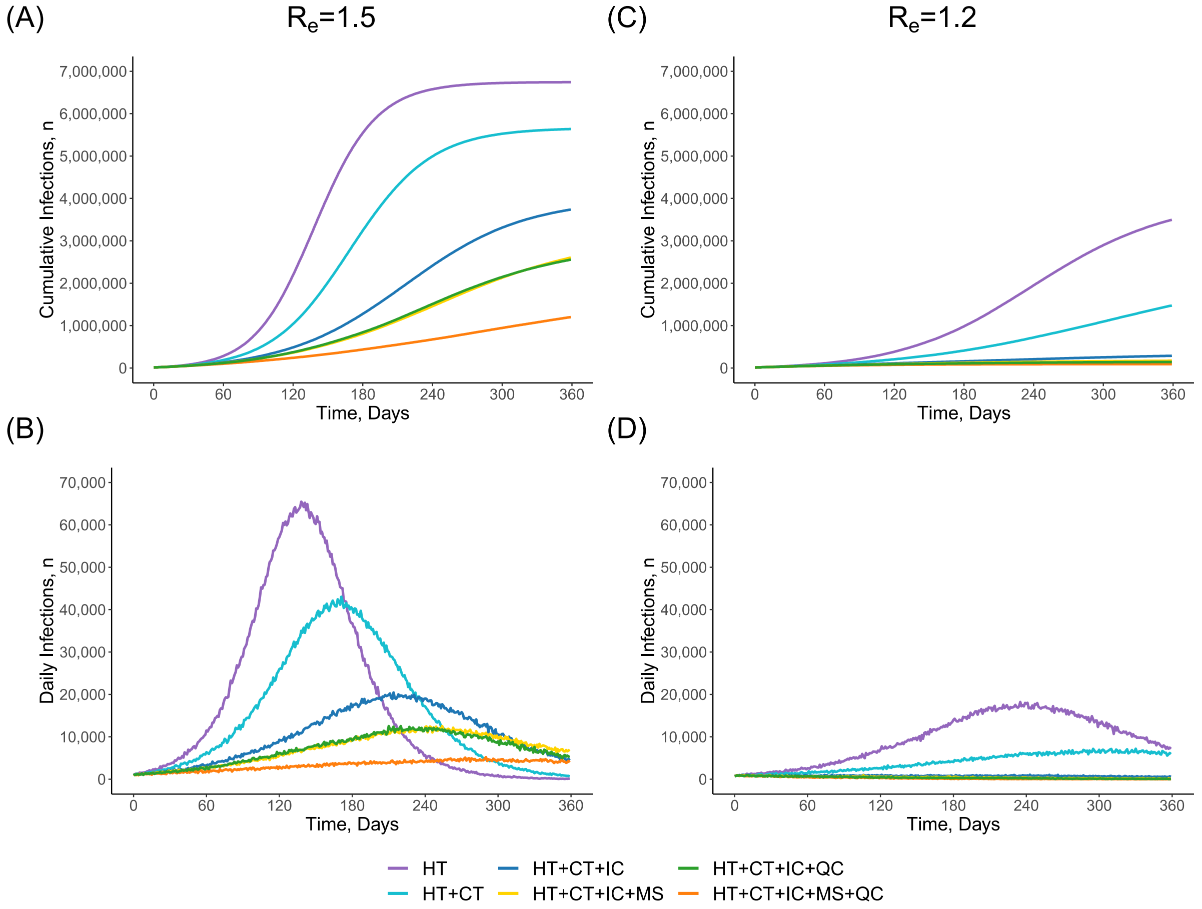


SARS-CoV-2: severe acute respiratory syndrome coronavirus 2. $R_e$: effective reproduction number. HT: healthcare testing. CT: contact tracing within households. IC: isolation centre. MS: mass symptom screen. QC: quarantine centre.

Panels A and B show model results with $R_e$=1.5. Panels C and D show model results with $R_e$=1.2. Panels A and C depict cumulative SARS-CoV-2 infections (both detected and undetected) over time by intervention strategy. Panels B and D depict daily SARS-CoV-2 infections. In each panel, time 0 on the horizontal axis represents the start of model simulation, with SARS-CoV-2 infection prevalence of 0.1% (~11,000 individuals with SARS-CoV-2 infection in KwaZulu-Natal, South Africa).

**Figure S4. Model-projected cumulative and daily COVID-19 deaths by intervention strategy in KwaZulu-Natal, South Africa.**

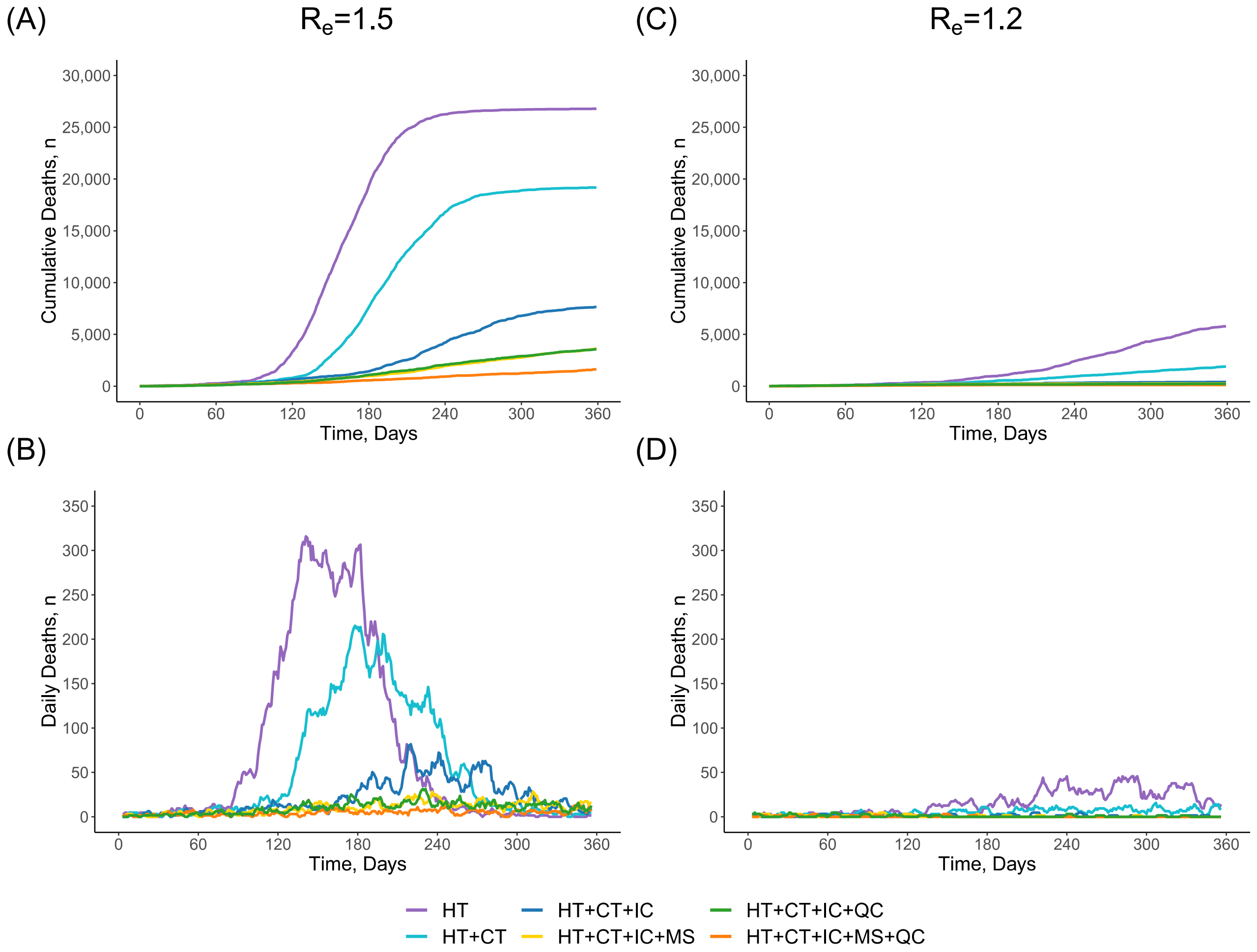


COVID-19: coronavirus diseases 2019. $R_e$: effective reproduction number. HT: healthcare testing. CT: contact tracing within households. IC: isolation centre. MS: mass symptom screen. QC: quarantine centre.

Panels A and B show model results with $R_e$=1.5. Panels C and D show model results with $R_e$=1.2. Panels A and C depict cumulative COVID-19 deaths over time by intervention strategy. Panels B and D depict 7-day averages of daily deaths due to COVID-19. In each panel, time 0 on the horizontal axis represents the start of model simulation, with SARS-CoV-2 infection prevalence of 0.1% (~11,000 individuals with SARS-CoV-2 infection in KwaZulu-Natal, South Africa).

**Figure S5. Multi-way sensitivity analysis demonstrating cost-effectiveness of strategies across a range of assumptions about the efficacies and costs of key public health interventions.**

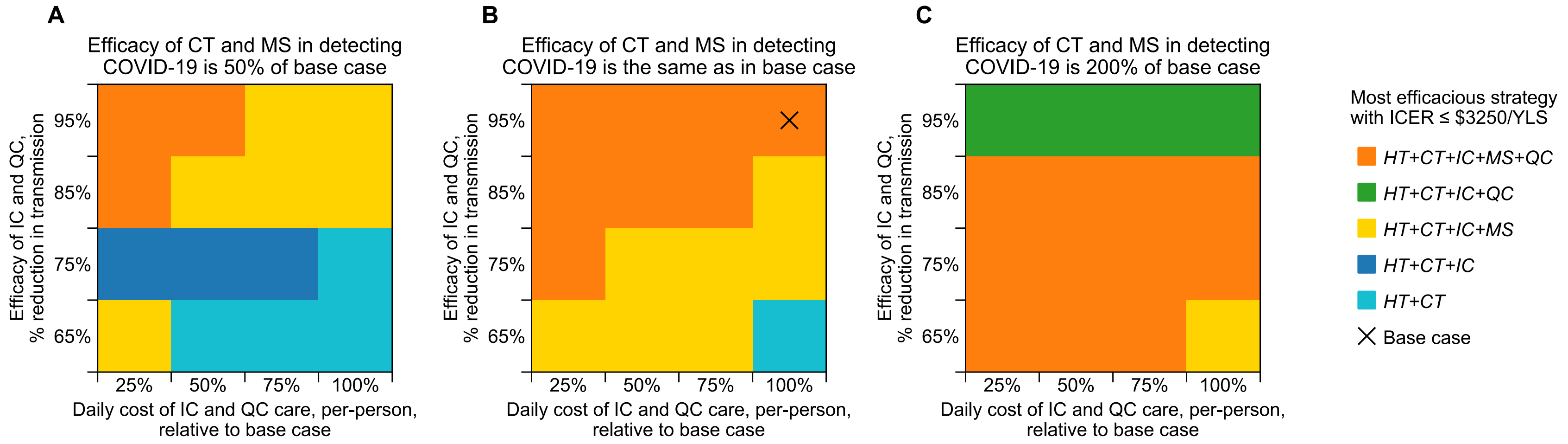


HT: healthcare testing. CT: contact tracing within households. IC: isolation centre. MS: mass symptom screen. QC: quarantine centre. ICER: incremental cost-effectiveness ratio. YLS: year-of-life saved.

The figure shows results of a multi-way sensitivity analysis in which we varied CT/MS efficacy in detecting COVID-19 cases, IC/QC efficacy in reducing transmission, and IC/QC costs. The color coding indicates the strategy that provided the greatest clinical benefit (YLS) while having an ICER that was below the cost-effectiveness threshold of $3,250/YLS.[1] In the base case, isolation centres and quarantine centres reduce transmission by 95%; isolation centre care costs $43·60/person/day and quarantine centre care costs $36·90/person/day. The effective reproduction number in this analysis was 1·5.

**Figure S6. Multi-way sensitivity analysis demonstrating cost-effectiveness of strategies across a range of assumptions about the efficacies and costs of key public health interventions, excluding quarantine centres as an option.**

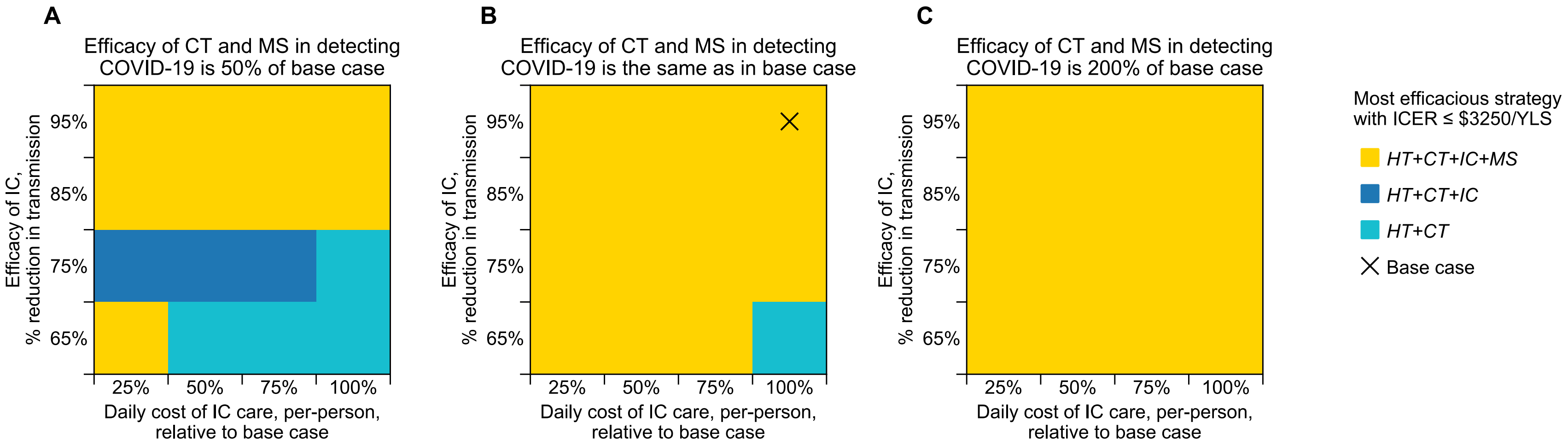


HT: healthcare testing. CT: contact tracing within households. IC: isolation centre. MS: mass symptom screen. ICER: incremental cost-effectiveness ratio. YLS: year-of-life saved.

The figure shows results of a multi-way sensitivity analysis in which we varied CT/MS efficacy in detecting COVID-19 cases, IC/QC efficacy in reducing transmission, and IC/QC costs. The color coding indicates the strategy that provided the greatest clinical benefit (YLS) while having an ICER that was below the cost-effectiveness threshold of $3,250/YLS.[1] In the base case, isolation centres and quarantine centres reduce transmission by 95%; isolation centre care costs $43·60/person/day and quarantine centre care costs $36·90/person/day. The effective reproduction number in this analysis was 1·5.
